# BrainPT, a Cognitive Mini-Game App for Short-Form Video Use in Young Adults: Neural and Behavioral Pilot Study

**DOI:** 10.64898/2026.10.11.26364666

**Authors:** Yunjeong Jeong, Chaewon Kim, Sungmin Ha, Seoyoung Choi, Sanghoon Han

**Affiliations:** Yonsei University

**Keywords:** short-form video, problematic digital media use, cognitive control, executive function, attention, electroencephalography, EEG, alpha rhythm, theta rhythm, functional connectivity, functional near-infrared spectroscopy, fNIRS, prefrontal cortex, dorsolateral prefrontal cortex, mobile health, mHealth app, behavior change, pilot study

## Abstract

**Background:** Short-form video elicits a stimulus-driven, passive neural state, yet existing interventions target time of use alone and cannot show whether the mechanism of cognitive control has changed.

**Objective:** This pilot study developed BrainPT, a mini-game-based cognitive intervention, and evaluated whether a single insertion alters the neural state during short-form viewing (Study 1) and whether repeated use improves behavioral self-regulation (Study 2).

**Methods:** Study 1 used a within-subject design (n=9) in which electroencephalography (EEG) and functional near-infrared spectroscopy (fNIRS) were recorded in the same session across short-form viewing, a single BrainPT set, and goal-directed tasks. Study 2 used a nonrandomized 2-week pretest-posttest design comparing an experimental group that used BrainPT daily (n=13) with a control group (n=8). Both visits followed the same in-laboratory protocol, and app logs and self-report indices were collected over the intervention period.

**Results:** In Study 1, short-form viewing was accompanied by prefrontal deactivation: EEG power fell below baseline in all four bands and most in alpha (−1.72 dB, *P*=.01; *q*=.054 after correction). Prefrontal activation was opposite in sign between the goal-directed tasks and viewing (+0.180 vs −0.093, *P*=.002), and the viewing block immediately after a BrainPT insertion exceeded the block before it (+0.110 vs −0.093, *P*=.02). The same condition ordering appeared in an independent preintervention sample (n=18). In Study 2, daily short-form use fell from 143.5 to 50.9 min in the experimental group and rose from 104.8 to 132.9 min in the control group, a between-group difference in change of 120.7 min (*P*=.004). The Adult ADHD Self-Report Scale did not differ between groups (*P*=.39), and no neural index separated the groups after 2 weeks.

**Conclusions:** A single insertion produced a measurable neural change: prefrontal state during the viewing that followed moved toward the goal-directed task. After 2 weeks of use that neural difference was not detectable between groups although behavior differed, so repeated use may act through the momentary state during viewing rather than by reorganizing the resting state. Both samples were small and nonrandomized, the primary outcome was self-reported, and the analyses were exploratory and not preregistered, so these estimates require confirmation in a preregistered randomized trial with objectively logged use.

**Trial Registration:** Clinical Research Information Service (CRIS), Republic of Korea, KCT0012762; https://cris.nih.go.kr

## Introduction

### Background

Short-form video moves fast. Clips are replaced within seconds, and the next one arrives faster than the last can be taken in. What comes next is chosen by a recommendation algorithm tuned to the individual, so the next clip is kept worth looking at. What distinguishes the format is not its content but its delivery structure: the stream itself supplies the reason to keep watching, so viewing requires neither a goal nor effort. Young adults are the heaviest users. Eighty percent of US adults aged 18-29 use Instagram and about half of that age group open TikTok at least once a day [1]. That is why this study targeted young adults.

### Prior Work

A systematic review and meta-analysis of short-form use reported a moderate association between heavier use and lower cognitive function (*r*=−.34), strongest for attention (*r*=−.38) and inhibitory control (*r*=−.41) [2]. Individual studies carry the association down to the neural level. Users with a stronger tendency toward short-form addiction showed weaker executive control on an attention-network task and a weaker frontal theta index of control [3], and shifted toward immediate reward in risky decision-making with prefrontal activation during that period varying by tendency [4]. Functional magnetic resonance imaging responses to personalized short-form video showed reduced self-regulation together with increased activity in the default mode network and the ventral tegmental area [5]. These studies established what state short-form use is associated with. Whether that state can be changed by an intervention is the next question, and it is where this study sits.

Intervention studies have mostly been evaluated with time of use and self-report. Restricting smartphone use improves stress, well-being, and sleep [6], but an umbrella review concluded that the effect estimates are highly heterogeneous and the certainty of evidence low [7]. One reason is that self-reported and logged use agree only moderately [8].

Yet one question remains open even with perfect measurement. Even if viewing time were logged exactly, "watching less" and "regulating better" would still not be distinguished. The two yield the same reduction in time but are different claims, and just as transfer beyond the trained task has rarely been confirmed in the cognitive training literature [9], a change in a behavioral index alone cannot specify what has changed. To adjudicate between them, the state the intervention is claimed to act on must be measured directly.

Targeting the state also fixes the sample and the indices. More adults report that their attention is fragmenting, but the diagnostic criteria for ADHD require several symptoms to have been present **before age 12** [10], so attentional change bound up with media encountered after that period falls outside the diagnostic category. That is why this study sampled general young adults rather than a diagnosed ADHD group, and measured ADHD tendency as a continuous scale (ASRS) rather than as a category.

### Short-Form Viewing as a Neural State

We therefore treat short-form viewing not as content consumption but as a neural state. When people watch edited video continuously, cortical activity synchronizes across viewers [11], and the magnitude of that synchronization varies with how much attention the viewer gives [12]. The brain state during viewing is measurable and is not fixed. Attention is organized into a goal-directed top-down system and a stimulus-driven bottom-up system that detects salient events and interrupts top-down control [13]. The short-form format continuously occupies the bottom-up system, so the state during viewing can be defined as a stimulus-reactive state, whose opposite pole is the goal-directed task state in which the prefrontal cortex maintains a goal representation [14]. The two states separate on measurable indices: alpha activity gates information flow and desynchronizes over engaged regions [15,16], frontal midline theta tracks control demand [17], and dorsolateral prefrontal activation rises during response inhibition and risky decision-making [18,19]. On this view the target of an intervention is not the total time of use but the state during viewing, and moving that state toward the control-dominant pole of the axis is where the intervention acts.

We developed BrainPT, a mini-game-based intervention aimed at this target. Rather than blocking viewing, BrainPT inserts a short cognitive task at the moment of consumption. The tasks borrow the formats of the Comprehensive Attention Test (CAT) [20] to engage subfunctions of attention and working memory, without administering the test itself as an assessment. There is precedent for game-format interventions changing neural indices of cognitive control: adaptive multitasking game training increased frontal midline theta power and frontal-posterior theta coherence, and the magnitude of that increase predicted improvement on an untrained sustained-attention task [21].

### Objectives and Hypotheses

If the target is a shift in neural state, two questions follow. First, does a single insertion immediately move the neural state during viewing in the intended direction? Second, when that immediate effect is repeated, is it accompanied by improved self-regulatory behavior? We examined these questions on two timescales. Study 1 recorded EEG and fNIRS within a single session to measure the neural state during viewing and the change around a BrainPT insertion (within-subject, two visits). Study 2 compared 2 weeks of at-home use against a control group on daily short-form viewing time and self-regulation indices, and measured neural change over the same period (controlled pretest-posttest). Because 2 weeks is short for claiming sustained neural change, Study 2 is framed as a 2-week take-home study whose primary outcome is behavioral, with neural carry-over examined as a secondary outcome without a prespecified direction.

The four hypotheses tested are given in Textbox 1.

#### Textbox 1

Four working hypotheses derived from theory. These were derived from anatomical and theoretical grounds rather than registered before data collection, and all analyses in this study are exploratory with respect to them as well.

*H1.* Short-form viewing elicits a stimulus-reactive neural state distinct from task engagement, with reduced prefrontal engagement (Study 1, neuroimaging).
*H2.* The difference between the two states is not explained by a single arousal axis (Study 1, neuroimaging).
*H3.* A cognitive task inserted at the moment of consumption moves the state along the control axis (Study 1, neuroimaging).
*H4.* A group that repeats that state shift for 2 weeks shows a larger reduction in daily short-form viewing time (Study 2, behavioral, nonrandomized).

## Methods

### Study Design

This report comprises two studies. Study 1 is a within-subject repeated-measures design with two laboratory visits and no app use between them; Study 2 is a nonequivalent control group pretest-posttest design with an unsupervised at-home use period between the two visits. The two studies were recruited separately and share no participants. Study 2 prospectively assigned participants to an intervention and therefore falls within the ICMJE definition of a clinical trial, which does not require randomization. It was not registered in a trial registry before enrollment; registration was not sought at the time because the design was nonrandomized. The study was registered retrospectively in the Clinical Research Information Service (CRIS), Republic of Korea, a WHO Primary Registry (KCT0012762), and the absence of prospective registration is stated as a limitation. Reporting follows the TREND statement for nonrandomized evaluations of behavioral interventions [22], and the intervention description follows CONSORT-EHEALTH [23] (completed checklists in Multimedia Appendices 4 and 5).

### Ethical Considerations

This study was conducted in accordance with the ethical principles of the Declaration of Helsinki and was approved by the Institutional Review Board of the Department of Psychology, Yonsei University (7001988-202509-HR-2369-04). Before starting, all participants were informed of the purpose, procedures, expected discomfort, scope of data use, and their right to withdraw at any time without penalty, and gave written consent. App usage records were transmitted to a private repository accessible only to the research team. Data were recorded with participant initials and a study number only; no direct identifiers such as names or contact details were stored, and initials will be replaced with random identifiers on release. Participants were compensated: KRW 30,000 (about US $22 at the exchange rate during the study period) for the two visits in Study 1, and in Study 2 KRW 30,000 for the control group and KRW 50,000 (about US $36) for the experimental group, who in addition to the two visits used the app and completed daily self-reports for about 2 weeks. The difference reflects that additional burden.

### Participants

Participants were recruited through online notices on a Korean university-community application. Inclusion criteria were university students in their 20s or 30s who watched short-form video for at least 30 minutes a day on average, wanted to reduce their own use, and did not expect a major change in phone use over the coming 2 weeks. Exclusion criteria were (1) a history of substance abuse or dependence, (2) current or past major psychiatric disorder (DSM-IV Axis I), (3) chronic medication that could alter neural patterns, and (4) uncorrected vision problems. Both sets of criteria were stated in the notice and confirmed in a pre-survey. Measurements took place in a university laboratory for about 1 hour per visit; recording periods were January-February 2025 for Study 1 and September-November 2025 for Study 2. Numbers at each stage and reasons for exclusion are given in Table 1.

**Table 1.** Participant characteristics, preintervention baseline values, and intervention exposure.

| Measure | Study 1 | Study 2 control | Study 2 experimental | Study 2 between-group $P^a$ |
| --- | --- | --- | --- | --- |
| **Sample<sup>c</sup>** |  |  |  |  |
| Enrolled or assigned, n | 15 | 8 | 15 | — |
| Behavioral analysis, n | — | 8 | 13 | — |
| Neural analysis — EEG, n | 9 | 8 | 10 | — |
| Neural analysis — fNIRS, n | 9 | 7 | 9 | — |
| **Demographics<sup>b</sup>** |  |  |  |  |
| Age (years), mean (SD; range) | 22.1 (1.9; 20–26) | 24.3 (3.5; 20–32) | 22.8 (2.3; 20–28) | .31 |
| Sex (male:female), n | 2:7 | 4:4 | 6:7 | >.99 |
| **Preintervention baseline** |  |  |  |  |
| Short-form use (min/day), mean (SD) | —<sup>c</sup> | 104.8 (37.6) | 143.5 (84.2) | .17 |
| ASRS total (18 items), mean (SD) | 33.7 (14.4) | 32.8 (14.7) | 32.1 (10.0) | .91 |
| **Intervention exposure<sup>d</sup>** |  |  |  |  |
| Total app trials, median (range) | — | 12 (12–13) | 372 (12–628) | — |
| Interval between visits (days), mean (SD)<sup>f</sup> | 22.1 (5.0) | 15.9 (3.0) | 19.8 (6.4) | .13 |
| Timing of measurement (days after study start), median | — | 44 | 10 | <.001 |

<sup>a</sup>Compares the Study 2 control and experimental groups. Sex, Fisher exact test; timing of measurement, Mann-Whitney U test; all others, Welch $t$ test.
<sup>b</sup>Each column uses its own analysis sample. Mean age in the experimental EEG sample ( $n=10$ ) was 22.2 years (SD 1.6) ( $P=.16$ ).
<sup>c</sup>Study 1 has no intervention period.
<sup>d</sup>All assigned participants.
<sup>e</sup>The interval between visits was computed only for participants with raw EEG at both visits ( $n=9/8/9$ ).
<sup>f</sup>Numbers excluded at each stage and the reasons are given in the Participants section.
ASRS: Adult ADHD Self-Report Scale; fNIRS: functional near-infrared spectroscopy.

Study 2 was not randomized. The two groups differed in compensation and participation conditions, were recruited through separate notices, and the experimental group was recruited and measured first, after which the control notice was posted. Groups are therefore defined by recruitment route and timing; applicants did not choose their condition, and neither participants nor experimenters were blinded. The control group completed daily self-reports for about 2 weeks without the app.

### Intervention (BrainPT)

BrainPT is an iOS application that, rather than blocking short-form consumption, inserts a short cognitive task at the point of use. Because iOS does not report short-form viewing time separately from app time, BrainPT uses daily cumulative use of the two applications through which short-form video is mainly consumed (Instagram and YouTube) as a proxy. When that time reaches the threshold (30 minutes for all participants), the app launches, presents one set of cognitive tasks, and closes when the set ends. Viewing is interrupted while the tasks are performed, but subsequent viewing was not blocked.

Ten tasks were implemented and five were presented per set. Borrowing CAT formats, the set targeted sustained attention, selective attention, interference control, divided attention, and working memory, and difficulty was fixed independently of performance. The app was distributed through iOS TestFlight; trial counts, accuracy, and completion time per game were uploaded automatically and used to verify adherence, and task performance itself was not treated as an outcome. The app is shown in Figure 1A and the screens and rules for all ten tasks in Multimedia Appendix 1.

**Figure 1.**
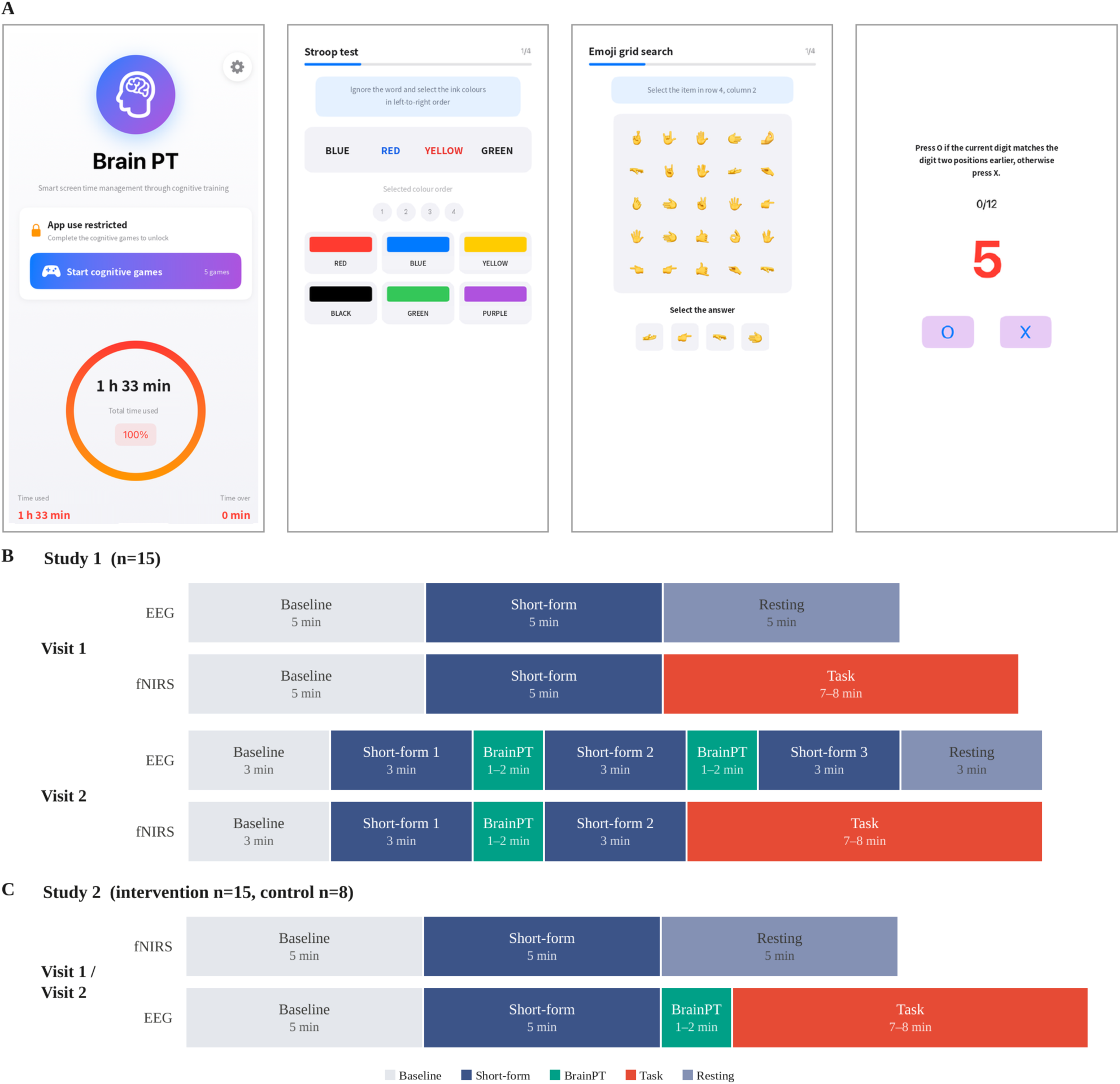
The BrainPT app and the experimental procedure. (A) From left, the home screen after the daily short-form threshold is reached and three cognitive tasks; the screens are shown translated into English, and the application used by participants was in Korean. Screens and rules for all ten tasks are given in Multimedia Appendix 1. (B) Study 1. (C) Study 2. The two visits share the same block structure and are shown once. Block width is proportional to duration; BrainPT and the cognitive tasks end on completion, so their lengths varied.

### Procedure

#### Study 1

Participants attended twice; at both visits EEG was recorded first and the equipment was then changed for fNIRS. Visit 1 was measured without an insertion and visit 2 with BrainPT inserted between short-form blocks (EEG twice, fNIRS once; Figure 1B). The Study 1 EEG had no task block, so the viewing-task contrast was derived from fNIRS. There was no app use between visits.

#### Study 2

Study 2 proceeded as visit 1, unsupervised at-home use, visit 2. The two visits applied the same protocol identically to both groups, and, opposite to Study 1, fNIRS was recorded first. Because the EEG contained both a BrainPT block and a task block, the viewing-task contrast and all BrainPT-related analyses were derived from EEG, while the fNIRS run comprised baseline, short-form, and resting blocks. Only the experimental group was instructed to install the app after visit 1, and both groups completed daily self-reports throughout the intervention period. The interval between visits was planned as 2 weeks, and the intervention period was defined identically for all participants as the 14 days immediately preceding visit 2.

### Measures

#### EEG

Recording used a 14-channel wireless Emotiv EPOC X at 128 Hz (AF3, F7, F3, FC5, T7, P7, O1, O2, P8, T8, FC6, F4, F8, AF4). The baseline block was defined as an eyes-open resting period.

#### fNIRS

Recording used a NIRSIT LITE at 8.138 Hz, with 15 standard channels and 6 short-separation channels. Short-separation channels were used to regress out systemic physiological components such as scalp blood flow. The primary region of interest was set on anatomical grounds to ch01 (Brodmann area [BA] 46, MNI coordinates 54.7/44.0/0.0).

#### Cognitive Tasks

Two laboratory tasks served as the goal-directed comparison condition: the Balloon Analogue Risk Task [24], in which a virtual balloon is inflated under increasing risk of loss, and an emotional Go/No-Go [25], which requires withholding a response to a designated emotional expression. The two tasks map onto risk-taking and response inhibition, core behavioral features of ADHD.

#### Questionnaires

Daily short-form viewing time (minutes) was derived by combining screen-time screenshots with a self-reported short-form proportion. ADHD tendency was measured with the 18-item Adult ADHD Self-Report Scale (ASRS), each item scored 0-4 (total 0-72) [26], with inattention (9 items) and hyperactivity-impulsivity (9 items) subscales. Age, sex, and habitual sleep duration were also collected. At visit 2 the experimental group additionally completed a 5-item post-use survey: three multiple-choice items on the effect of BrainPT on cognitive ability, the urge to watch, and anxiety when interrupted, and two open-ended items on the experience of use and desired improvements.

#### Daily Self-Report

Collected in Study 2 only. During the intervention period both groups rated five items daily from 1 to 5 ("not at all" to "very much"). The items covered that day’s sustained attention, emotional stability, impulse control, plan execution, and overall satisfaction (for example, the sustained-attention item was "Today I was able to see through a task that required concentration").

### Neuroimaging Data Processing

EEG was processed in the following order. A 1-40 Hz band-pass filter was applied, bad channels were detected and interpolated, and the data were re-referenced to the common average. Independent component analysis was then run and components classified by ICLabel as eye-blink, muscle, or noise with probability at least 0.80 were removed, and blocks were segmented on markers. Welch power spectra were estimated within each block (**1-s window, 50% overlap, 1 Hz resolution**) and finally normalized in dB against the baseline block of the same session.

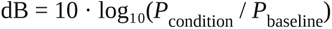

From these values we derived band power (delta 1-4, theta 4-8, alpha 8-12, beta 12-30 Hz) and channel-pair weighted phase lag index (wPLI) [27]. Coupling was computed for theta, alpha, and beta only, because 1-4 Hz phase estimation is unstable in short blocks. The Study 2 cognitive task block was truncated to the first 300 s because completion time varied across participants (the Study 1 EEG has no task block).

fNIRS was motion-corrected, converted to concentration with the modified Beer-Lambert law, passed through short-separation regression and drift control (0.005-0.1 Hz), segmented on markers, and fitted with a general linear model to obtain oxygenated hemoglobin (HbO) betas per condition. For both modalities the identical pipeline was applied before any group split. Step-by-step criteria and settings are given in Multimedia Appendix 2.

### Outcome Measures

Theory-based indices were defined on the following grounds. Frontoparietal coupling is the mean wPLI of four channel pairs (F3-P7, F4-P8, AF3-P7, AF4-P8) [28]; the high-resolution analysis in Study 1 uses channel F4; the fNIRS region of interest is ch01 (BA46). F4 and ch01 correspond to the same right prefrontal region. Because absolute power differs across participants, all analyses use values normalized to the same-session baseline.

Study 1 is a proof-of-concept study with no outcome hierarchy and examined four things: EEG band power and fNIRS ch01 HbO beta during the short-form block, the 0.5 Hz resolution analysis of the F4 power spectrum, the difference between short-form and cognitive task blocks, and the difference between the short-form blocks before and after a BrainPT insertion together with change across time-thirds of the viewing block. All are relative to the same-session baseline.

The **primary outcome of Study 2 is behavioral change after 2 weeks of use**: the change in daily short-form viewing time and the between-group difference in that change. ASRS total, app logs, and five laboratory cognitive-task indices were examined alongside. The **secondary outcome is neural carry-over**: the visit 1 to visit 2 change in frontoparietal coupling, global mean coupling, band power, and fNIRS ch01, compared between groups with no direction specified in advance. Two analyses that use visit 1 data only — the replication of the Study 1 condition contrasts and the condition structure of frontoparietal coupling — address replication rather than intervention effects and sit outside this hierarchy. Exhaustive connectivity exploration, dose-response by baseline use, and the post-use survey items are reported as exploratory indices.

### Statistical Analysis

Indices are reported as **theory-based** (a small set defined on anatomical and theoretical grounds) or **exhaustive** (all possible indices computed and then compared), a distinction based on the grounds for selecting the index. Effect sizes are *d*_z_ for paired comparisons and Cohen *d* for between-group comparisons, and multiplicity was controlled with the Benjamini-Hochberg false discovery rate.

#### Study 1

Condition contrasts were judged with paired *t* tests. Because adjacent frequency bins are not independent, the high-resolution analysis used cluster-based correction, with the primary test an exhaustive sign-flip test enumerating all 2⁸=256 sign patterns for 8 participants and an AR(1) null distribution reported alongside. Change across viewing segments was tested with an exact paired permutation test enumerating the full 2⁹=512 permutation space for 9 participants.

#### Study 2

Intervention effects were judged with change-score *t* tests together with analysis of covariance using visit 1 as the covariate (V2 ∼ group + V1), and were not accepted when the two disagreed. A within-group test significant in only one group was not used as evidence of a between-group difference [29]. Between-group comparisons used Welch *t* tests and Welch intervals because dispersion differed, and absence of effect was addressed with equivalence testing (TOST) [30]. Behavioral outcomes are reported with as-assigned and per-protocol analyses and with a sensitivity analysis restricted to the neural analysis sample. Remaining procedures and electrode-level values are given in Multimedia Appendices 2 and 3.

## Results

### Study 1

#### The Neural State During Short-Form Viewing

##### During short-form viewing, power in all four bands shifted below baseline (Figure 2A)

Averaged across the 14 channels and expressed in dB against the same-session baseline, the decrease was largest in alpha (−1.72 dB, 95% CI −2.97 to −0.46, *P*=.01, *d*_z_=−1.05; 8 of 9 participants), followed by theta, delta, and beta (−1.11, −0.76, −0.63). Only alpha was significant before correction, and applying Benjamini-Hochberg false discovery rate control across the four bands left it just short of threshold (*q*=.054).

**Figure 2.**
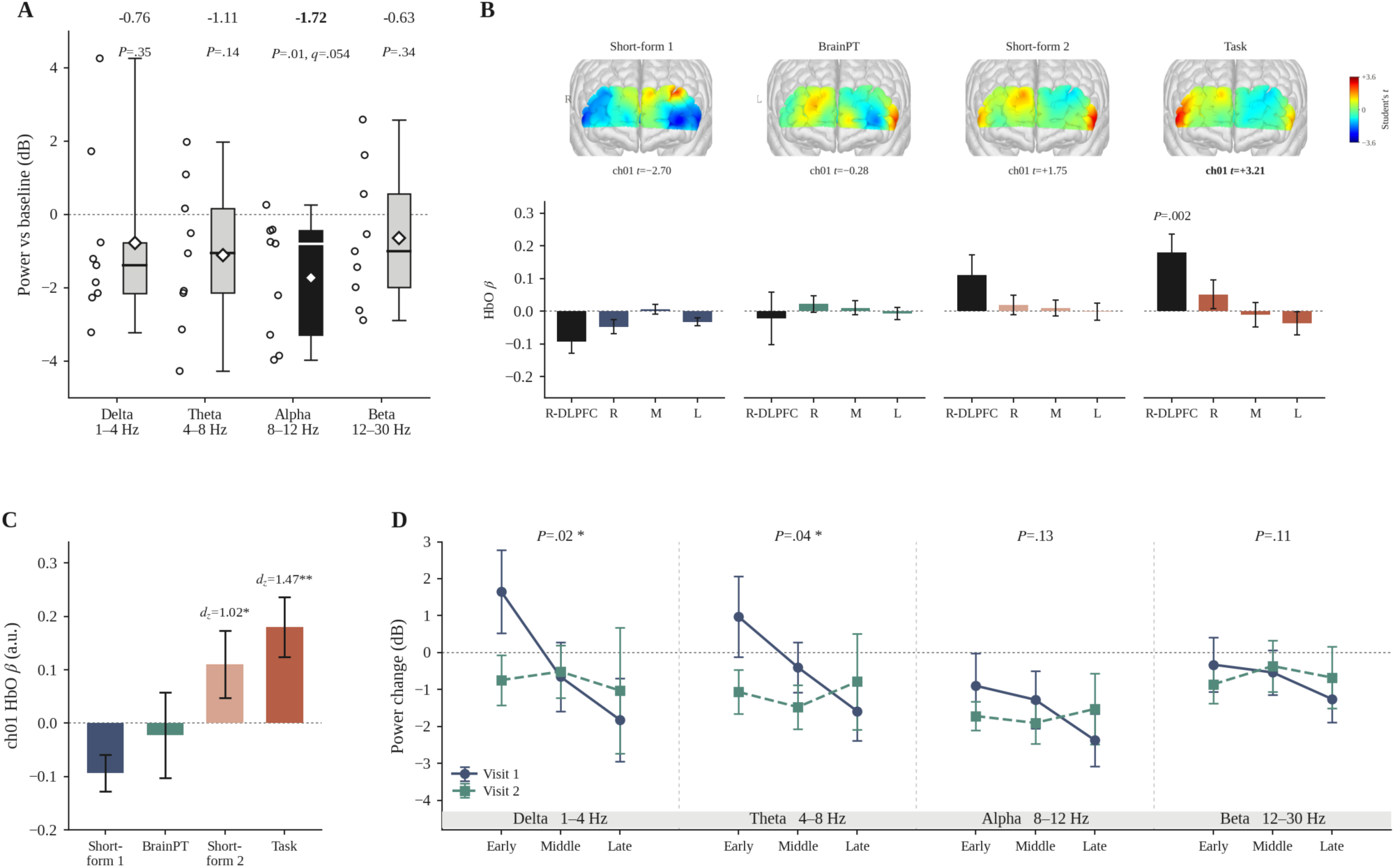
The neural state during short-form viewing and the effect of the inserted intervention (Study 1, n=9). (A) EEG band power during short-form viewing, averaged over 14 channels, in dB relative to baseline. Open circles are individual participants, boxes show the interquartile range and median, whiskers the observed range, and white diamonds the mean; only bands significant before correction are filled. (B) Prefrontal oxygenated hemoglobin across the visit 2 block sequence. Above, the map (color is Student *t* against baseline, ±3.6); below, values by location for the region of interest ch01 (right dorsolateral) and the right, middle, and left groupings, with the *P* shown for the short-form 1 versus task comparison. The left-right partition is an approximation based on channel-coordinate correspondence. (C) Block-by-block values at ch01 and effect sizes for the paired comparison against short-form 1. (D) Change in band power across viewing segments: visit 1 is the three time-thirds of a single block and visit 2 the three blocks separated by BrainPT. Error bars are the standard error of the mean and *P* is the exact permutation value for the visit × segment interaction.

##### In the fNIRS of the same session, activation of the right dorsolateral prefrontal cortex (ch01, BA46) was also below rest

(HbO GLM β=−0.093, 95% CI −0.173 to −0.014; Figure 2C). This decrease came from the component that moves with the task rather than from the background component of the signal. The intercept and exponent of the aperiodic component [31] stayed at the same level across the two conditions (Δ=−0.072, *P*=.62; Δ=+0.039, *P*=.73; n=8), and the activity of the components removed by ICA [32] likewise did not vary with condition (Δ=−0.044, *P*=.31) and was independent of the aperiodic change (*r*=0.10, 0.11; both analyses use visit 1 data, where short-form viewing is a single block). The electrophysiological index (EEG band power) and the hemodynamic index (fNIRS prefrontal activation) both fell below rest within the same session.

#### Frequency Specificity of the Decrease

##### Before correction, the decrease was concentrated in low alpha (8.5-10.5 Hz) rather than spread across the alpha band

When the F4 power spectrum relative to baseline was computed at 0.5 Hz resolution (1-30 Hz, 59 bins, n=8), the decrease was significant in 11 bins, and the longest contiguous run among them was 8.5-10.5 Hz (cluster mass 17.42). This cluster did not survive correction for multiple comparisons (*P*=.09; AR(1) null distribution *P*=.06). No other frequency region showed a significant decrease even before correction. At the channel level the strongest decreases were alpha at F4 and beta at AF4 and FC6, all three in the right frontal region (all uncorrected).

#### Contrast With the Cognitive Tasks

##### Activation in the same channel was below rest during short-form viewing and above rest during the cognitive tasks (Figure 2C)

HbO beta during the cognitive tasks (Balloon Analogue Risk Task and emotional Go/No-Go) was +0.180 (95% CI 0.051 to 0.309), placing it on the opposite side of rest from short-form viewing (−0.093). The difference between the two conditions survived false discovery rate correction and ran in the same direction in 8 of 9 participants (Δβ=+0.273, *P*=.002, *q*=.009, *d*_z_=1.47). Of the two comparison tasks, Go/No-Go has right frontal activation related to response inhibition summarized at the meta-analytic level [18], and engagement of the same region has also been reported for risky decision-making [19].

#### The Effect of the Inserted Intervention

##### With BrainPT placed in between, the short-form block that followed rose to a state close to that of the task

The visit 2 fNIRS run was ordered short-form, BrainPT, short-form, task (Figure 2B and 2C). The short-form block before the insertion was below rest (β=−0.093), whereas the short-form block after it rose above rest and approached the task level (+0.180) at β=+0.110. The shift between the two short-form blocks survived false discovery rate correction and ran in the same direction in 8 of 9 participants (*P*=.02, *q*=.03, *d*_z_=1.02). The post-insertion short-form block was not itself separable from rest (*P*=.12). Activation during the BrainPT block was not distinguishable from the short-form block immediately before it (*P*=.32) and differed from the task (*P*=.04). The shift therefore appeared not in the BrainPT block itself but in the short-form block that followed it.

##### In the EEG, the decline over viewing time was markedly attenuated when an insertion was present (Figure 2D)

At visit 1, without an insertion, dividing the viewing block into thirds showed all four bands falling toward later segments, with a first-to-last difference of 3.47 dB in delta. At visit 2, with BrainPT inserted, the same span was only 0.28 dB and alpha even rose slightly. The visit × segment interaction was nominally significant for delta and theta (*P*=.02, .04) but not for alpha or beta, and no band survived false discovery rate correction across the four.

### Study 2

The results below are presented in the order of replication of the Study 1 condition contrasts, behavioral change after 2 weeks of use, and neural carry-over while the app is not in use.

#### Replication of the Study 1 Findings

##### The between-condition state differences observed in Study 1 replicated in a nonoverlapping independent sample

All Study 2 participants underwent the same measurements at the preintervention visit 1, so these data test replication rather than an intervention effect, with twice the sample (n=18) and a task block that the Study 1 EEG did not have. Task block data were obtained in 17 of 18 participants, so all short-form-task comparisons are n=17.

Band power by condition was lowest during short-form viewing, intermediate during BrainPT, and highest during the cognitive task (Figure 3A). During viewing the mean was below baseline in all four bands and during the cognitive task above baseline in all four, and the short-form-task difference was significant for delta, theta, and alpha (*P*=.018, .002, .035; beta *P*=. 064).

**Figure 3.**
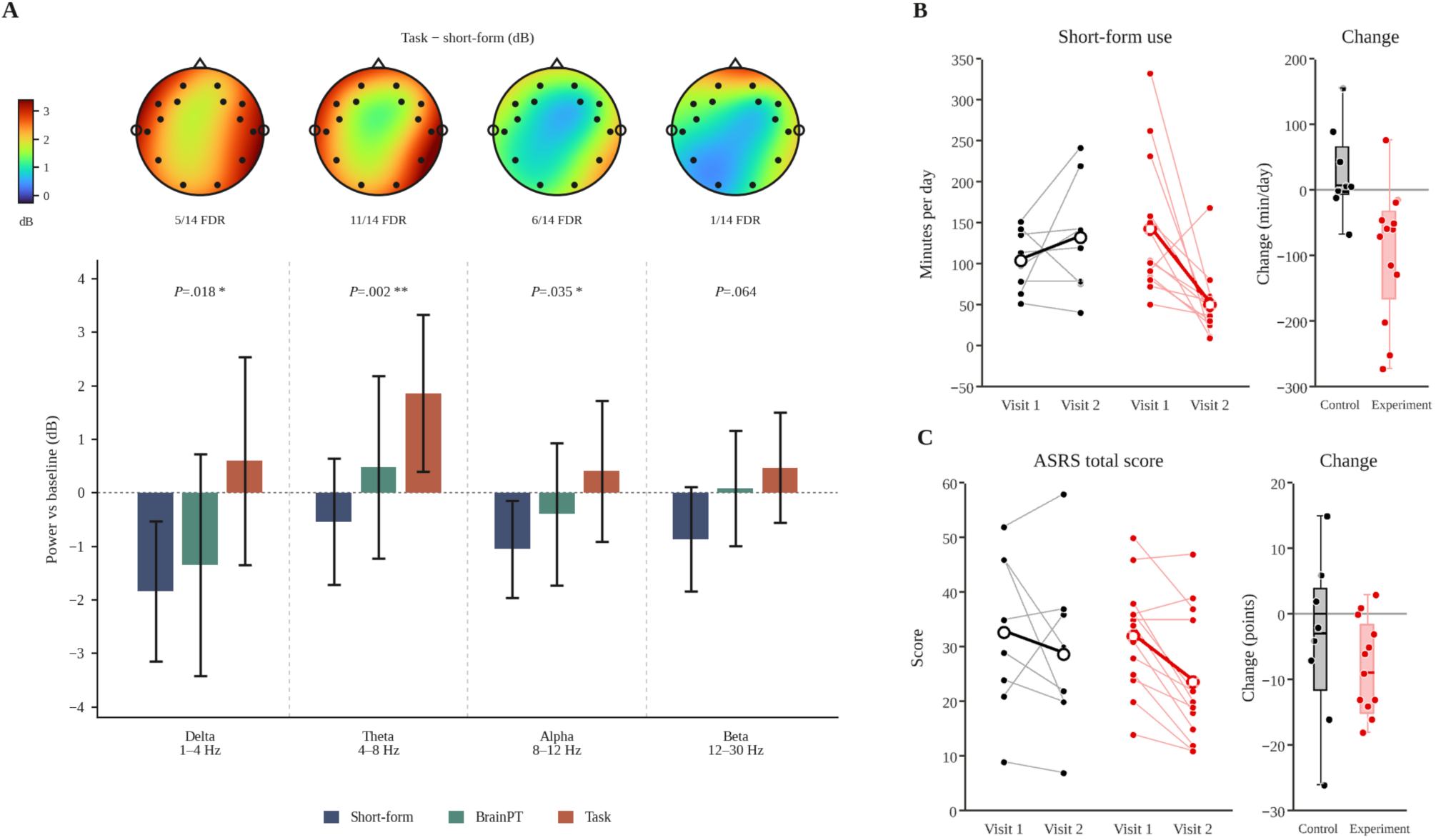
Neural state by condition and behavioral change after 2 weeks in Study 2. (A) EEG band power by condition, averaged over 14 channels, in dB relative to the same-session baseline (visit 1, n=18; cognitive task n=17). Bars are group means and vertical lines 95% CIs; the *P* shown is the uncorrected paired short-form-task comparison (one asterisk *P*<.05, two *P*<.01). The topographies above are task minus short-form, with the number of channels surviving false discovery rate correction noted below. This montage has no midline or central electrodes, so the interior of the head is interpolated. (B) Daily short-form viewing time and (C) Adult ADHD Self-Report Scale total, the behavioral indices after 2 weeks of use. Control is black and experimental red. In each left panel thin lines are individual participants and the thick line with open markers is the group mean. Each right panel shows per-participant change as points with the interquartile range and median as a box. Between-group comparisons of change are given in the text.

The contrasts agreed in direction across the two modalities. In the Study 1 fNIRS the right dorsolateral prefrontal cortex (BA46) moved from −0.093 during viewing to +0.180 during the cognitive task (*d*_z_=1.47, *P*=.002, n=9; Figure 2B), and in the Study 2 EEG theta power across the same three conditions was −0.54, +0.49, and +1.86 dB (Figure 3A). The shift was confined to the region of interest: in the frontal pole (BA10) of the same fNIRS recording there was no difference between viewing and task (Δ=+0.020, *P*=.54).

#### Behavioral Change After 2 Weeks of Use

##### After 2 weeks, daily short-form viewing in the experimental group fell by 65% while the control group did not decrease (Figure 3B)

The two groups did not differ significantly in short-form viewing time or ASRS total before the intervention, but baseline viewing time in the experimental group was numerically higher and more variable (143.5, SD 84.2 vs 104.8, SD 37.6 min/day; Table 1), so results are reported as change from each participant’s own baseline.

Daily short-form viewing in the experimental group fell from 143.5 to 50.9 minutes, a decrease of 92.5 minutes (12 of 13 participants, *P*=.006, *d*_z_=0.93), while the control group rose from 104.8 to 132.9 minutes, which was not significant (+28.1, *P*=.28). The between-group difference in change was significant (Δ=−120.7 min, 95% CI −197.4 to −43.9, *P*=.004, *d*=−1.35). Analysis of covariance with visit 1 as covariate gave the same conclusion (group coefficient −81.6 min, *P*=.005), and a sensitivity analysis restricted to the neural data sample preserved both direction and significance (*P*=.02).

##### ASRS total decreased in both groups and the between-group difference did not reach significance (Figure 3C)

The experimental group fell from 32.1 to 23.7 (−8.4, *P*=.001; 10 of 13 participants) and the control group from 32.8 to 28.8 (−4.0, *P*=.40). The decrease was larger in the experimental group (Δ=−4.4, 95% CI −15.4 to 6.6, *P*=.39, *d*=−0.46; analysis of covariance agreed, *P*=.31). There were also no between-group differences on the five laboratory cognitive-task indices (*P*=. 09-.97). Exploratorily, splitting the experimental group at its preintervention median (105 min) showed a much larger decrease in the higher-use subgroup (−156.3 min, n=7 vs −18.2 min, n=6; *P*=.005).

Adherence was verified from app logs. Excluding one nonadherent participant (12 sessions), use in the experimental group was 57-628 sessions (median 374) over a median of 13 days, while the control group’s 12-13 sessions were in-laboratory exposure. In the post-use survey all 13 experimental participants reported that the insertion helped restrain viewing, and some wrote that they reduced viewing because the task was bothersome rather than because their state had changed.

#### The Condition Structure of Frontoparietal Coupling

##### Frontoparietal coupling showed the same ordering as band power: lowest during short-form viewing and higher during BrainPT (Figure 4A)

wPLI across the four theory-based pairs (F3-P7, F4-P8, AF3-P7, AF4-P8) fell below baseline during viewing and rose above baseline during BrainPT in all three bands. The magnitude of change was similar across bands (Δ=+0.063 to +0.075) but most consistent in beta, where all 18 participants moved in the same direction and the change exceeded twice the between-participant variability (*d*_z_=2.31 vs 1.09, 1.08; *q*<.001). At the coupling level BrainPT exceeded not only viewing but also the cognitive task, which differs from power, where BrainPT lay between the two conditions.

**Figure 4.**
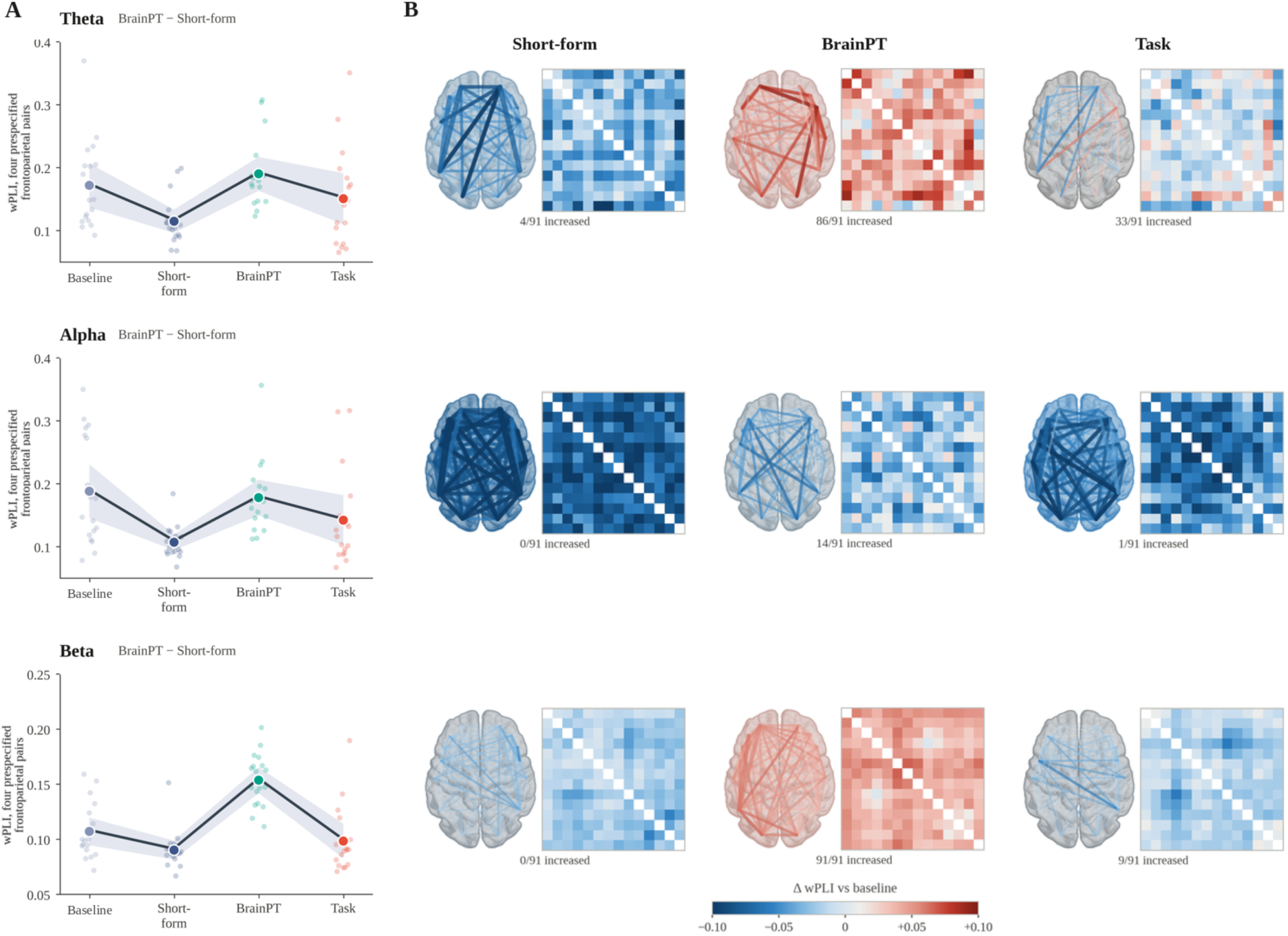
Scalp coupling by condition (Study 2 visit 1, n=18; cognitive task n=17). (A) Weighted phase lag index for the four theory-based frontoparietal pairs across baseline and the three conditions. Next to each band title is the BrainPT-short-form contrast tested in the text; the difference and effect size are given in the text. (B) Scalp coupling by condition across all 91 channel pairs; rows follow the same band order as (A) and columns the conditions in session order. Each cell shows a scalp network and the 14×14 channel-pair matrix of the same data, with color the change in weighted phase lag index relative to baseline, clipped at ±0.10. Networks are viewed from above with anterior upward, and the bottom row gives the number of pairs that increased. Data are in sensor space and are not source estimates.

##### This rise was not confined to the four theory-based pairs but extended across the scalp (Figure 4B)

Values below are relative to baseline. Across all 91 channel pairs, beta coupling during BrainPT was above baseline in all 91 pairs and theta in 86. In the same bands the numbers of pairs that increased during viewing were 0 and 4, and during the cognitive task 9 and 33. Alpha decreased in most pairs in all three conditions (BrainPT 14, viewing 0, task 1), opposite to the other two bands.

#### Neural Carry-Over

##### After 2 weeks of repeated use, no neural index measured while the app was not in use separated the two groups

The criterion was between-group analysis of covariance (V2 ∼ group + V1). Band power changed similarly in all four bands (*P*=. 37-.93), and right dorsolateral prefrontal HbO rose more in the experimental group but not significantly (+0.095 vs +0.014, *P*=.36). Beta coupling in the BrainPT condition for the theory-based frontoparietal index also increased more in the experimental group without reaching significance (Δ=+0.034 vs +0.010; 9 of 10 participants, *P*=.09), the 91-pair global mean coupling showed the same pattern (Δ=+0.026 vs +0.008, *P*=.07), and the remaining conditions were similar (*P*=.22-.75).

Beyond the theory-based indices, 504 between-group analyses of covariance were run on all-channel band power, all-region-pair coupling, all-channel fNIRS, hemodynamic response shape, and signal entropy, and none survived correction for multiple comparisons (minimum *q*=.11). Restricting to the 17 indices that met the test-retest reliability criterion (*r*≥0.5) gave the same result. Change-score tests agreed with the analysis of covariance except for one index. In equivalence testing, 10 of 12 indices met equivalence at this sample’s minimum detectable effect (*d*=1.42) and none met it at the conventional bound (*d*=0.5).

#### Exploratory Connectivity Analysis

The following is an exhaustive exploration without correction for multiple comparisons. For each of the 12 cells formed by three bands × four conditions, 31 coupling indices were computed (the mean over all 91 pairs, 14 per-channel node strengths, the means of the 15 region pairs formed by five regions, and the theory-based frontoparietal mean) and compared between groups by analysis of covariance with visit 1 as covariate (372 tests in total). The question examined was whether significant results were spread evenly across the 12 cells or concentrated in one.

Fifteen were significant, which does not exceed chance expectation (about 19). However, 10 of them fell in the single beta × BrainPT cell (about 1.6 expected per cell), and in that cell all 31 indices favored the experimental group. All 10 significant indices were posterior, and the theory-based frontoparietal mean was not significant in this cell (*d*=0.60-0.96). Because the most concentrated of 12 cells was selected, however, the concentration is not significant once cell selection is corrected for (*P*=.35).

The same condition was also examined at the channel-pair level. Among the 91 pairs in the beta BrainPT condition, 17 were significant and 16 of those increased more in the experimental group (uncorrected *P*<.05; about 4.6 expected by chance), but the four theory-based frontoparietal pairs were not among them (*P*=.09-.39).

## Discussion

### The Neural State During Short-Form Viewing

In the results, power in all four EEG bands fell below baseline during short-form viewing, and in the fNIRS of the same session right dorsolateral prefrontal activation was also below rest. That two indices with different measurement principles pointed in the same direction is what this section has to explain.

The decrease is not explained by reduced arousal. Lower arousal should raise low-frequency power, and if arousal were a single axis, a low-frequency decrease should be accompanied by a beta increase; instead all four bands fell together. Taken alone, the conventional reading of an alpha decrease is release from inhibition, that is, that the region was engaged by the task [15,16]; but frontal theta, which tracks control demand [17], and prefrontal blood flow moved in the same direction, so it cannot be read as increased engagement either. What remains is that the state during viewing is not a low-arousal state but one in which the prefrontal control system is not engaged [14].

The decrease was not spread evenly across the spectrum but concentrated in low alpha (8.5-10.5 Hz) and in right frontal channels. What these two coordinates mean is that the EEG decrease points to the same place as the fNIRS decrease. The channels with the largest decrease were alpha at F4 and beta at AF4 and FC6, all three right frontal, and the fNIRS decrease was likewise in the right dorsolateral prefrontal cortex. Two instruments with different principles indicated the same region. This frequency concentration did not survive cluster correction (*P*=.09), however, so it does not carry the weight of the two preceding lines of evidence.

This account fits the literature on naturalistic video viewing. Edited video synchronizes cortical activity across viewers [11] and the magnitude of that synchronization varies with how much attention the viewer gives [12], which means that viewing is stimulus-driven processing rather than passive exposure. Attention divides into a goal-directed and a stimulus-driven system, and salient stimuli interrupt the former [13]; short-form is a format that triggers that interruption repeatedly on a short cycle. What this study observed is the state in which the goal-maintaining system stands back while that format occupies attention.

### Contrast With the Cognitive Tasks

In the results, activation in the same fNIRS channel was below rest during short-form viewing and above rest during the cognitive tasks, so the three conditions separated with rest in between. If short-form viewing were merely a low-demand cognitive activity, its activation should lie between rest and the cognitive task; this arrangement is therefore explained by a difference in kind rather than in degree of engagement.

Of the two comparison tasks, Go/No-Go has right frontal activation related to response inhibition summarized at the meta-analytic level [18], and engagement of the same region has also been reported for risky decision-making [19]. This rise indicates that the region is in a state of maintaining a goal and selecting a response [14]. That the same region falls below rest during viewing suggests a state in which the control system is not engaged. The two conditions separate not in the magnitude of prefrontal engagement but in its direction relative to rest, and that axis defines the target of the intervention.

### The Effect of the Inserted Intervention

In the results, a single BrainPT insertion raised prefrontal activation in the immediately following viewing block to a level close to the task condition, and also attenuated the decline over time within the block. The key to the interpretation is that the shift appeared not in the BrainPT block itself but in the viewing block right after it. That the prefrontal cortex engages while a task is being performed is unremarkable; that the state carried into the viewing that followed the task leads to the account that the insertion briefly changed the mode of processing of viewing itself.

The larger shift in frontoparietal coupling points the same way. This circuit has been described as the route by which anterior regions drive posterior ones when top-down control prevails over bottom-up signals [28], and that coupling during BrainPT exceeded even the level of the cognitive task fits the account that the insertion strongly, if temporarily, recruited the control-side configuration. Coupling is a scalp-level index, however, so a change in the circuit itself cannot be asserted. That the state moves not by blocking viewing but by inserting a different demand into its stream is the mechanistic core of this study.

### Replication in an Independent Sample

In the results, the preintervention data of Study 2 reproduced the between-condition arrangement of Study 1. This replication is more than a repetition because the test conditions differed. The sample does not overlap and is twice as large, and it included a task block that the Study 1 EEG lacked, so the viewing-task contrast could be confirmed directly in EEG. In Study 1 this contrast rested on a single modality, fNIRS; obtaining the same direction in a different modality and a different sample makes it hard to attribute the observed arrangement to a property of a particular instrument or sample.

### Behavioral Change After 2 Weeks of Use

In the results, daily short-form viewing in the experimental group fell by 65% after 2 weeks of at-home use, with no significant change in the control group. Participants were not told to cut down, and the reduction was far larger in those with higher preintervention use.

This dose-response pattern is consistent with the intervention working as designed. The condition under which the intervention fires is the moment daily cumulative use crosses the threshold, so heavier users encounter more firing points and correspondingly more room for change. What was targeted was not overall use habits but consumption beyond the threshold, and that is exactly where the change appeared.

The scope of the change also matches that target. ASRS and the laboratory cognitive tasks were similar before and after, so what is supported here is behavioral change in the targeted consumption context; improvement in attentional function in general would require a separate test.

### The Condition Structure Seen in Coupling

In the results, frontoparietal coupling at visit 1 showed the same ordering as band power. That the arrangement of lowest during short-form viewing and highest during BrainPT appeared in both indices means that it is not a property of one measurement method.

The point where the two indices diverge is the BrainPT condition. In power, BrainPT lies between viewing and the cognitive task, whereas in coupling it exceeded the cognitive task, and the rise extended beyond the four theory-based pairs across the scalp. Power measures the magnitude of local recruitment and coupling the phase alignment between separated sites, so the two indices show different faces. That the inserted task falls short of the laboratory cognitive task in total local recruitment while exceeding it in between-region alignment is the observation obtained here, and it is a clue to how a brief insertion recruits the control-side configuration.

That alpha moved opposite to theta and beta in all three conditions also fits the conventional reading of alpha. Alpha has been associated with disengaging regions that are not involved rather than binding those that are [15,16], so a decrease in every condition away from baseline is compatible with the rise in the other two bands.

This arrangement is a between-condition contrast at a single time point, visit 1, so between-group comparisons are treated separately in the two sections that follow.

### Neural Carry-Over

In the results, no neural index measured after 2 weeks of use while the app was not running separated the two groups. This intervention works at the level of the state during use, and repeating it is accompanied by an actual reduction in use behavior, but no evidence was obtained that repetition also changed the state while the app was not in use.

This null result does not conflict with the neuroplasticity literature on cognitive training. Training studies that reported neural-level change are generally longer and more intensive. Adaptive multitasking game training changed frontal midline theta and frontal-posterior coupling in 12 prescribed hours of training over 4 weeks [21]. What this study observed is a few minutes a day of voluntary use over 2 weeks, so the result is better explained by placing the intervention on a dose axis. How much state-level effect must accumulate before it crosses into a change in the resting state is not yet settled.

An exploration sweeping all indices likewise found no index separating the groups, although uncorrected significant results were concentrated in a single cell, beta coupling in the BrainPT condition. That cell is the task condition the experimental group alone repeated for 2 weeks, so the concentration can be read either as a neural effect of the intervention or as task familiarity. Because it is not significant once cell selection is corrected for, no direction is taken here and it is left as a target for a prespecified test.

### The Role of BrainPT

This study did not set out to demonstrate the efficacy of the app; it used the app as a means of testing whether the principle of inserting an intervention at the moment of consumption holds. The observed behavioral change is therefore better attributed to the whole process of becoming aware of one’s own use time and performing a different action on the spot than to the performance of the app itself. Participants’ accounts are consistent with this, but they also point to a second route: several reduced viewing not because their state during viewing changed but because the inserted task was bothersome and they wanted to avoid it. Avoiding the interruption and shifting the state during use produce the same reduction in viewing time and are not separable in this design. The two routes do make different predictions, however. If avoidance is the main route, the effect should weaken as the task becomes familiar; if state shift is the main route, the quality of viewing immediately after an insertion should differ. The latter is the direction indicated by the single-insertion result of this study, but whether that direction holds over 2 weeks of use was not confirmed.

### Comparison With Prior Work

The relation between short-form use and attention has so far been reported as correlations between amount of use and self-report or task performance, or as comparisons between groups high and low in use tendency [2–5]. What this study adds is a move of the unit of observation from individual differences to state. What was observed here is not that heavier users are more distractible but that within the same person prefrontal engagement during viewing falls below rest and deepens as viewing continues; and unlike an individual-difference variable, a state variable can be acted on in the moment.

The difference from digital-use interventions lies in the target. Existing interventions mostly target the total directly, through blocking or time limits [6,7]. This intervention did not target the total and did not instruct participants to cut down, yet the total fell in the group that used it. Blocking interventions hold their effect while the blocking is maintained, whereas here the total fell while use continued, and adherence was maintained throughout the 2-week logs. This reduction cannot be separated from the between-group difference in timing of measurement, however, and there was no active control, so the comparison remains at the level of a literature contrast.

The absence of far transfer repeatedly reported in the cognitive training literature [9] was reproduced here as well: there was no intervention effect on the laboratory cognitive tasks. Even so, change did appear in an everyday use behavior whose form is entirely unlike the trained tasks. This suggests that whether transfer succeeds may depend on target setting more than on training dose. Targeting general cognitive ability does not carry beyond the trained task, but targeting the state of a specific consumption context changed behavior in that same context. If this distinction holds, the important question in designing digital well-being interventions is closer to where one trains than to what one trains.

### Limitations

#### Nonrandomized Assignment

The two groups were not randomly assigned, and because the experimental group was recruited and measured first, timing of measurement differed between groups (median day 10 vs day 44), so intervention effects and timing effects are not separable. All 5 participants excluded from the neural data were in the experimental group (5/15 vs 0/8, Fisher exact test *P*=.12).

#### Sample and Power

Because group samples are small (experimental n=13-15, control n=8), the neural null results may reflect insufficient power. The minimum effect detectable at 80% power is *d*=1.42, and detecting *d*=0.5 would require about 64 participants per group.

#### Measurement

Short-form viewing time relied on self-report and submitted screen-time screenshots, and ASRS is also self-report, so the principal behavioral indices are all participant-reported. The intervention’s firing threshold likewise used Instagram and YouTube app time as a proxy rather than short-form viewing time.

#### Neural Measurement

The EEG is 14-channel sensor-space data, so the coupling reported is a scalp-level index and falls short of the recommended minimum for sensor-level connectivity (32 channels). The fNIRS covered only 15 prefrontal channels, and because the channel coordinates are not published the spatial representation is an approximation. Because the channel removed under the bad-channel cap rule varies between runs in preprocessing, the absolute band-power values in Study 2 are one realization; the ordering across conditions was preserved in all eight implementations (Multimedia Appendix 2). Task completion time fell between visits (median 400.6 s to 357.9 s), so the cognitive task condition was not identical at the two time points.

#### Absence of Preregistration and Trial Registration

This study was not preregistered and the analysis plan was not fixed in writing before the data were inspected. Study 2 was also not registered in a trial registry before enrollment, although prospective assignment to an intervention brings it within the ICMJE definition of a clinical trial; it was registered retrospectively in the Clinical Research Information Service (CRIS), Republic of Korea (KCT0012762), and the absence of prospective registration removes one safeguard against selective reporting. The *P* values reported are not corrected over the space of all analyses performed, so the estimates obtained here serve to generate hypotheses.

#### Exploratory Finding

Outside the theory-based indices, change was concentrated in posterior beta-band BrainPT coupling (10 of 15; permutation *P*=.052), but the location is not the a priori frontoparietal one and the BrainPT condition is a task the experimental group alone repeated, so state change and task familiarity cannot be distinguished. It is not taken as evidence of an intervention effect.

#### Duration

Two weeks may be short for observing state-level change from repeated use. This study is a 2-week take-home study rather than a chronic intervention study.

### Conclusions

This exploratory pilot study showed that short-form viewing can be defined as a specific neural state and that an insertion-type intervention aimed at that state is associated with change in the state and in subsequent use. Prefrontal engagement during viewing was below rest, and it was distinguished from active cognitive tasks by the direction of prefrontal control rather than by level of arousal. Inserting a cognitive task once without breaking the viewing stream moved this state toward the cognitive-task pole, and after 2 weeks of repeated at-home use daily short-form viewing fell by 65% (*d*=−1.35). That the total fell without being targeted shows that intervention designs not reliant on blocking or time limits are possible. What this study did not show is equally clear: at the 2-week point the two groups did not separate on any neural index. The present evidence therefore reaches only the claim that this intervention works at the level of the state during use, not the claim that repetition reorganizes the resting state, and it was obtained under the constraints of nonrandomized assignment and small samples.

## Supporting information

Multimedia_Appendix_1_Task_screens

Multimedia_Appendix_2_Methods_detail

Multimedia_Appendix_3_Electrode_table

Multimedia_Appendix_4_TREND_checklist

Multimedia_Appendix_5_CONSORT_EHEALTH

## Data Availability

All data produced in the present study are available upon reasonable request to the authors. Raw electroencephalography and functional near-infrared spectroscopy recordings are not deposited in a public repository because they contain potentially identifiable signal characteristics. Analysis code is publicly available at https://github.com/rachelkiim/brainpt.

## Acknowledgments

We thank the participants for their time. We thank Seojin Kim, Sumin Lee, and Seoyoung Jang for their contributions to discussions during the study design phase, and CogSci:IN, the cognitive science society of Yonsei University, which provided the setting in which the intervention concept was first developed. No generative artificial intelligence tools were used in producing the scientific content, data, or analyses reported in this manuscript.

## Data Availability

The data generated and analyzed in this study are available from the corresponding author on reasonable request. Analysis code is publicly available [33]. Raw electroencephalography and functional near-infrared spectroscopy data are not deposited in a public repository because they contain potentially identifiable signal characteristics.

## Authors’ Contributions

YJ and CK conceptualized the study. YJ, CK, SH, and SC prepared the experimental protocol, conducted the experimental sessions, and analyzed the data. The BrainPT application was developed by the authors, with implementation led by SH. YJ and CK wrote the first draft. SHa supervised the study and reviewed and edited the manuscript. All authors read and approved the final manuscript.

## Abbreviations

ADHD: attention-deficit/hyperactivity disorder
ASRS: Adult ADHD Self-Report Scale
BA: Brodmann area
CAT: Comprehensive Attention Test
EEG: electroencephalography
fNIRS: functional near-infrared spectroscopy
GLM: general linear model
HbO: oxygenated hemoglobin
ICA: independent component analysis
MNI: Montreal Neurological Institute
SEM: standard error of the mean
wPLI: weighted phase lag index

## References

1. Gottfried, J., & Park, E. (2025, November 20). Americans’ social media use 2025. Pew Research Center. https://www.pewresearch.org/internet/2025/11/20/americans-social-media-use-2025/

2. Nguyen, L., Walters, J., Paul, S., Monreal Ijurco, S., Rainey, G. E., Parekh, N., Blair, G., & Darrah, M. (2025). Feeds, feelings, and focus: A systematic review and meta-analysis examining the cognitive and mental health correlates of short-form video use. Psychological Bulletin, 151(9), 1125–1146. 10.1037/bul0000498

3. Yan, T., Su, C., Xue, W., Hu, Y., & Zhou, H. (2024). Mobile phone short video use negatively impacts attention functions: An EEG study. Frontiers in Human Neuroscience, 18, 1383913. 10.3389/fnhum.2024.1383913

4. Zhang, S., & Li, S. (2025). How short video addiction affects risk decision-making behavior in college students based on fNIRS technology. Frontiers in Human Neuroscience, 19, 1542271. 10.3389/fnhum.2025.1542271

5. Su, C., Zhou, H., Gong, L., Teng, B., Geng, F., & Hu, Y. (2021). Viewing personalized video clips recommended by TikTok activates default mode network and ventral tegmental area. NeuroImage, 237, 118136. 10.1016/j.neuroimage.2021.118136

6. Pieh, C., Humer, E., Hoenigl, A., Schwab, J., Mayerhofer, D., Dale, R., & Haider, K. (2025). Smartphone screen time reduction improves mental health: A randomized controlled trial. BMC Medicine, 23(1), 107. 10.1186/s12916-025-03944-z

7. Lu, P., Qiu, J., Huang, S., Wang, X., Han, S., Zhu, S., Ning, Y., Zeng, F.-F., & Yuan, Y. (2025). Interventions for digital addiction: Umbrella review of meta-analyses. Journal of Medical Internet Research, 27, e59656. 10.2196/59656

8. Parry, D. A., Davidson, B. I., Sewall, C. J. R., Fisher, J. T., Mieczkowski, H., & Quintana, D. S. (2021). A systematic review and meta-analysis of discrepancies between logged and self-reported digital media use. Nature Human Behaviour, 5(11), 1535–1547. 10.1038/s41562-021-01117-5

9. Simons, D. J., Boot, W. R., Charness, N., Gathercole, S. E., Chabris, C. F., Hambrick, D. Z., & Stine-Morrow, E. A. L. (2016). Do “brain-training” programs work? Psychological Science in the Public Interest, 17(3), 103–186. 10.1177/1529100616661983

10. American Psychiatric Association. (2022). Diagnostic and statistical manual of mental disorders (5th ed., text rev.). American Psychiatric Association Publishing.

11. Hasson, U., Nir, Y., Levy, I., Fuhrmann, G., & Malach, R. (2004). Intersubject synchronization of cortical activity during natural vision. Science, 303(5664), 1634–1640. 10.1126/science.1089506

12. Ki, J. J., Kelly, S. P., & Parra, L. C. (2016). Attention strongly modulates reliability of neural responses to naturalistic narrative stimuli. The Journal of Neuroscience, 36(10), 3092–3101. 10.1523/JNEUROSCI.2942-15.2016

13. Corbetta, M., & Shulman, G. L. (2002). Control of goal-directed and stimulus-driven attention in the brain. Nature Reviews Neuroscience, 3(3), 201–215. 10.1038/nrn755

14. Miller, E. K., & Cohen, J. D. (2001). An integrative theory of prefrontal cortex function. Annual Review of Neuroscience, 24, 167–202. 10.1146/annurev.neuro.24.1.167

15. Klimesch, W., Sauseng, P., & Hanslmayr, S. (2007). EEG alpha oscillations: The inhibition-timing hypothesis. Brain Research Reviews, 53(1), 63–88. 10.1016/j.brainresrev.2006.06.003

16. Jensen, O., & Mazaheri, A. (2010). Shaping functional architecture by oscillatory alpha activity: Gating by inhibition. Frontiers in Human Neuroscience, 4, 186. 10.3389/fnhum.2010.00186

17. Cavanagh, J. F., & Frank, M. J. (2014). Frontal theta as a mechanism for cognitive control. Trends in Cognitive Sciences, 18(8), 414–421. 10.1016/j.tics.2014.04.012

18. Simmonds, D. J., Pekar, J. J., & Mostofsky, S. H. (2008). Meta-analysis of Go/No-go tasks demonstrating that fMRI activation associated with response inhibition is task-dependent. Neuropsychologia, 46(1), 224–232. 10.1016/j.neuropsychologia.2007.07.015

19. Cazzell, M., Li, L., Lin, Z.-J., Patel, S. J., & Liu, H. (2012). Comparison of neural correlates of risk decision making between genders: An exploratory fNIRS study of the Balloon Analogue Risk Task (BART). NeuroImage, 62(3), 1896–1911. 10.1016/j.neuroimage.2012.05.030

20. Yoo, H. K., Lee, J.-S., Kang, S.-H., Park, E.-H., Jung, J.-S., Kim, B.-N., Son, J.-W., Park, T.-W., Kim, B.-S., & Lee, Y.-S. (2009). Standardization of the Comprehensive Attention Test for the Korean children and adolescents. Journal of the Korean Academy of Child and Adolescent Psychiatry, 20(2), 68–75.

21. Anguera, J. A., Boccanfuso, J., Rintoul, J. L., Al-Hashimi, O., Faraji, F., Janowich, J., Kong, E., Larraburo, Y., Rolle, C., Johnston, E., & Gazzaley, A. (2013). Video game training enhances cognitive control in older adults. Nature, 501(7465), 97–101. 10.1038/nature12486

22. Des Jarlais, D. C., Lyles, C., & Crepaz, N. (2004). Improving the reporting quality of nonrandomized evaluations of behavioral and public health interventions: The TREND statement. American Journal of Public Health, 94(3), 361–366. 10.2105/ajph.94.3.361

23. Eysenbach, G. (2011). CONSORT-EHEALTH: Improving and standardizing evaluation reports of web-based and mobile health interventions. Journal of Medical Internet Research, 13(4), e126. 10.2196/jmir.1923

24. Lejuez, C. W., Read, J. P., Kahler, C. W., Richards, J. B., Ramsey, S. E., Stuart, G. L., Strong, D. R., & Brown, R. A. (2002). Evaluation of a behavioral measure of risk taking: The Balloon Analogue Risk Task (BART). Journal of Experimental Psychology: Applied, 8(2), 75–84. 10.1037/1076-898X.8.2.75

25. Hare, T. A., Tottenham, N., Davidson, M. C., Glover, G. H., & Casey, B. J. (2005). Contributions of amygdala and striatal activity in emotion regulation. Biological Psychiatry, 57(6), 624–632. 10.1016/j.biopsych.2004.12.038

26. Kessler, R. C., Adler, L., Ames, M., Demler, O., Faraone, S., Hiripi, E., Howes, M. J., Jin, R., Secnik, K., Spencer, T., Ustun, T. B., & Walters, E. E. (2005). The World Health Organization adult ADHD self-report scale (ASRS): A short screening scale for use in the general population. Psychological Medicine, 35(2), 245–256. 10.1017/S0033291704002892

27. Vinck, M., Oostenveld, R., van Wingerden, M., Battaglia, F., & Pennartz, C. M. A. (2011). An improved index of phase-synchronization for electrophysiological data in the presence of volume-conduction, noise and sample-size bias. NeuroImage, 55(4), 1548–1565. 10.1016/j.neuroimage.2011.01.055

28. Buschman, T. J., & Miller, E. K. (2007). Top-down versus bottom-up control of attention in the prefrontal and posterior parietal cortices. Science, 315(5820), 1860–1862. 10.1126/science.1138071

29. Nieuwenhuis, S., Forstmann, B. U., & Wagenmakers, E.-J. (2011). Erroneous analyses of interactions in neuroscience: A problem of significance. Nature Neuroscience, 14(9), 1105–1107. 10.1038/nn.2886

30. Lakens, D. (2017). Equivalence tests: A practical primer for t tests, correlations, and meta-analyses. Social Psychological and Personality Science, 8(4), 355–362. 10.1177/1948550617697177

31. Donoghue, T., Haller, M., Peterson, E. J., Varma, P., Sebastian, P., Gao, R., Noto, T., Lara, A. H., Wallis, J. D., Knight, R. T., Shestyuk, A., & Voytek, B. (2020). Parameterizing neural power spectra into periodic and aperiodic components. Nature Neuroscience, 23(12), 1655–1665. 10.1038/s41593-020-00744-x

32. Muthukumaraswamy, S. D. (2013). High-frequency brain activity and muscle artifacts in MEG/EEG: A review and recommendations. Frontiers in Human Neuroscience, 7, 138. 10.3389/fnhum.2013.00138

33. Jeong, Y., Kim, C., Ha, S., Choi, S., & Han, S. (2026). BrainPT: Analysis code for a study of short-form video use [Computer software]. GitHub. https://github.com/rachelkiim/brainpt

