## Supplementary material for "BrainPT, a Cognitive Mini-Game App for Short-Form Video Use in Young Adults: Neural and Behavioral Pilot Study": Multimedia_Appendix_1_Task_screens

### Multimedia Appendix 1. Screens and rules for the 10 BrainPT cognitive tasks

Ten tasks were implemented in the BrainPT application. Each time the daily short-form threshold was reached, one set of five tasks was presented; difficulty was fixed and did not adapt to performance. Task formats were borrowed from the Comprehensive Attention Test to engage the intended functions; the test itself was not administered as an assessment. Trial count, accuracy, and response time were logged per task and uploaded automatically. The application interface used by participants was in Korean; the screens shown below are translated into English.

#### Task list

| # | Task | Intended function | Rule |
|---|---|---|---|
| 1 | Stroop | Interference control | Four colour words are shown, each printed in an incongruent ink colour. The participant selects the ink colours in left-to-right order, ignoring the words. |
| 2 | Emoji grid search | Selective attention | A 5×5 grid of similar hand emoji is shown. The participant reports the item at a stated row and column from four alternatives. |
| 3 | Letter grid search | Selective attention | A 5×5 grid of letters is shown. The participant reports the letter at a stated row and column from four alternatives. |
| 4 | Shape counting | Sustained attention | A 6×6 grid of coloured shapes is shown. The participant counts how many match a target colour–shape combination and types the number. |
| 5 | Aperture direction | Selective attention | A 5×5 grid of squares, each open on one side, is shown. The participant reports the opening direction of the square at a stated row and column. |
| 6 | Shape–colour matching | Divided attention | Two figures are shown. The participant responds by direction according to whether shape, colour, both, or neither match. |
| 7 | 2-back digits | Working memory | Digits appear one at a time. The participant indicates whether the current digit matches the one two positions earlier (12 trials). |
| 8 | Direction sequence: encoding | Working memory | A sequence of four directions is displayed for study. |
| 9 | Direction sequence: recall | Working memory | The participant reproduces the studied sequence by pressing direction keys in order. |
| 10 | Spatial order recall | Working memory | Cells of a 7×7 grid turn green one at a time. The participant then presses the cells' numbers in the order they changed. |

#### Screens

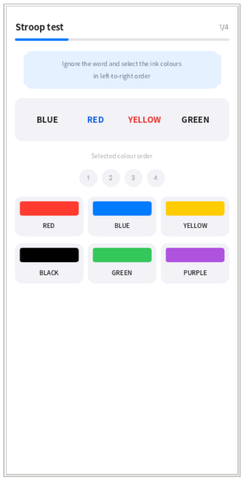


1. Stroop
interference control

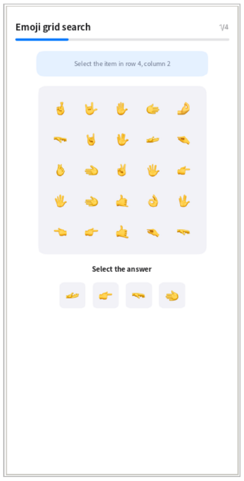


2. Emoji grid search
selective attention

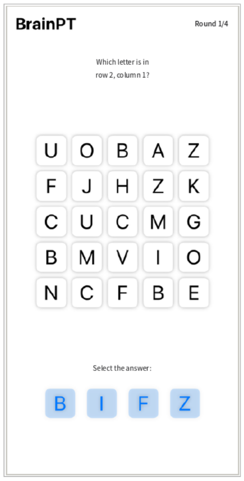


3. Letter grid search
selective attention

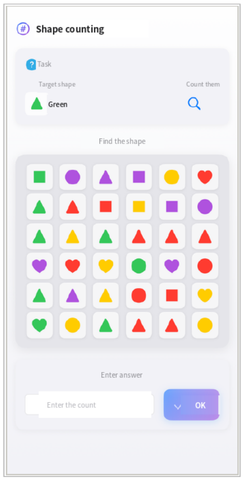


4. Shape counting
sustained attention

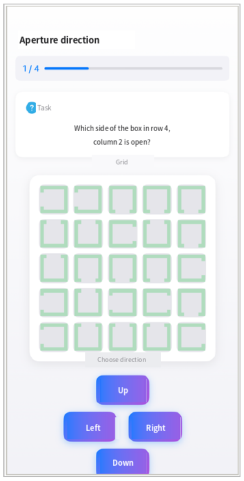


5. Aperture direction
selective attention

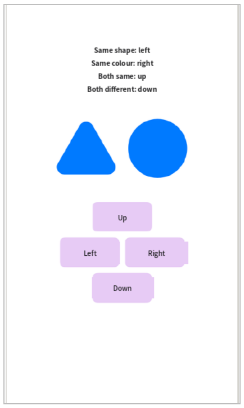


6. Shape–colour matching
divided attention

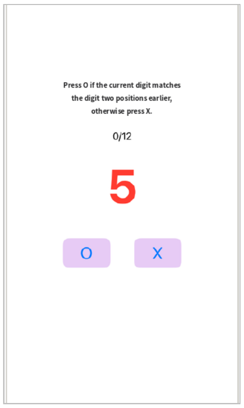


7. 2-back digits
working memory

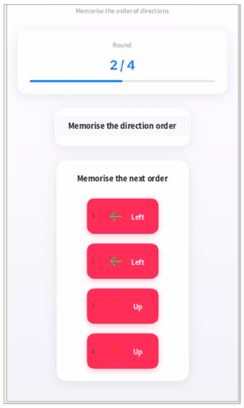


8. Direction sequence: encoding
working memory

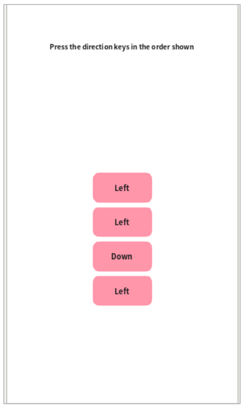


9. Direction sequence: recall
working memory

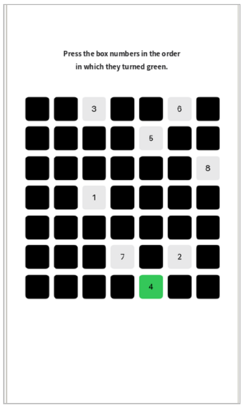


10. Spatial order recall
working memory

Screens are shown in the order of the table above. Numbers in the upper right of a screen indicate progress within the set.
