## Supplementary material for "BrainPT, a Cognitive Mini-Game App for Short-Form Video Use in Young Adults: Neural and Behavioral Pilot Study": Multimedia_Appendix_2_Methods_detail

### Multimedia Appendix 2. Segment matching, regional block contrasts, reliability criteria, and preprocessing settings

**1. Segment matching for the time-course analysis (Study 1).** The visit 1 and visit 2 protocols differ in how short-form viewing is partitioned, so the two visits cannot be compared block for block. At visit 1 viewing was a single uninterrupted 5-minute block; at visit 2 the same total viewing time was delivered as three blocks of about 3 minutes separated by two BrainPT insertions. To place the two visits on one axis, the visit 1 block was divided into three equal time-thirds and each third was matched to the visit 2 block occupying the same ordinal position: early to short-form 1, middle to short-form 2, late to short-form 3. The comparison is therefore of position within viewing, not of clock time, and the visit 2 segments carry the additional property of having been preceded by an insertion. Power spectra were computed separately within each segment and expressed in dB relative to the same-session baseline block, so segment length does not enter the contrast. Because the visit 1 thirds are contiguous while the visit 2 blocks are separated by the insertion, a visit × segment interaction cannot by itself distinguish a genuine effect of the insertion from an effect of interrupting a block; this is stated as a limitation in the main text.

**2. Regional block contrasts (Study 2).** The 14 electrodes were grouped into five scalp regions (frontal, fronto-central, temporal, parietal, occipital), giving 15 region pairs. For every region pair, frequency band (theta, alpha, beta) and condition (baseline, short-form, BrainPT, cognitive task), weighted phase lag index was compared between the experimental and control groups by analysis of covariance with the visit 1 value as a covariate, giving 180 models. Nine models reached $P<.05$ uncorrected, which is close to the number expected by chance; none survived Benjamini-Hochberg correction within the family (smallest $q$=0.78). The regional analysis therefore agrees with the channel-pair analysis reported in the main text. It is reported here rather than in the main text because the region grouping is a display convenience rather than an anatomical claim: these are sensor-space data and a region label describes where an electrode sits on the scalp, not which cortical area generated the signal.

**3. Reliability criteria for Study 2.** Indices entered the group comparison only if they were measurable at both visits under the same configuration, and three criteria were applied before any group contrast. First, a participant contributed an index only if the corresponding block existed and passed preprocessing at both visits; participants failing this at visit 2 were excluded from that index rather than imputed, which is why the analyzable sample differs across indices and is stated for each. Second, blocks whose recorded length departed from the protocol were not analyzed, because dB values referenced to a same-session baseline are stable only when the compared segments are of comparable length; short-form blocks were 306 (SD 13) seconds at the analyzed visit and BrainPT blocks 64 (SD 18) seconds, the latter varying because the task set ends on completion. Third, because the visit 1 measurement doubles as the replication sample for the Study 1 state findings, the visit 1 data were analyzed once for that purpose and once as the covariate in the intervention analysis; no index was selected for the intervention analysis on the basis of its visit 1 result.

**4. Preprocessing criteria and settings.** EEG was band-pass filtered at 1-40 Hz, bad channels were detected and interpolated, and the data were re-referenced to the common average. Bad-channel detection used a single channel-correlation criterion, and a cap rule removes only a subset when the number of flagged channels exceeds a limit. After independent component analysis, components classified by ICLabel as eye-blink, muscle, or noise with probability at least 0.80 were removed. Blocks were segmented on markers and Welch power spectra estimated per block (1-s window, 50 percent overlap, 1 Hz resolution); only the high-resolution F4 analysis in Study 1 used a 2-s window at 0.5 Hz resolution. The Study 2 cognitive task block was truncated to the first 300 s; actual task lengths were 320.9-600.5 s at visit 1 and 308.7-436.3 s at visit 2, so no participant fell below 300 s. fNIRS was motion-corrected, converted to concentration with the modified Beer-Lambert law, and cleaned of systemic physiology by short-separation regression. Drift was controlled in the 0.005-0.1 Hz band, blocks were segmented on markers, and a general linear model was fitted to obtain HbO betas per condition. Both modalities were normalized against the baseline block of the same session, and the identical pipeline was applied before any group split.

**5. Run-to-run variation of the bad-channel cap rule.** When the cap is active, the combination of channels removed varies between runs. Running the same code on the same data three times moved the 12 condition × band values of Study 2 visit 1 by 0.38 dB on average and 0.84 dB at most (the cap was active in 2 of 9 participants in Study 1 and 14 of 18 in Study 2). The ordering across conditions was preserved in all eight implementations for all four bands, and the short-form-task contrast came out stronger than the main-text value in every rerun. In the Study 2 neural carry-over analysis, a subset of 17 indices whose visit 1 to visit 2 correlation was $r \geq 0.5$ was reported alongside as a sensitivity analysis.
