## Supplementary material for "BrainPT, a Cognitive Mini-Game App for Short-Form Video Use in Young Adults: Neural and Behavioral Pilot Study": Multimedia_Appendix_3_Electrode_table

### Multimedia Appendix 3. Electrode-level band power by condition

Group mean (SD) power change in dB relative to the same-session baseline block, Study 2 visit 1. Short-form and BrainPT are based on 18 participants and the cognitive task on 17 (one participant had no task block). These are the values underlying the 14-channel mean plotted for each condition in Figure 3A. Electrodes are ordered in left-right pairs from frontal to occipital. These are sensor-space data; an electrode label indicates a scalp position, not a cortical source.

| | Delta | | | Theta | | | Alpha | | | Beta | | |
|---|---|---|---|---|---|---|---|---|---|---|---|---|
| **Electrode** | Short-form | BrainPT | Task | Short-form | BrainPT | Task | Short-form | BrainPT | Task | Short-form | BrainPT | Task |
| AF3 | -3.39 (4.41) | -2.00 (4.41) | -0.73 (5.66) | -1.60 (3.62) | 0.11 (4.04) | 1.03 (4.23) | -1.65 (2.34) | -0.53 (3.54) | 0.01 (3.74) | -1.84 (3.44) | -0.69 (3.40) | 0.39 (2.16) |
| AF4 | -2.91 (4.66) | -2.85 (5.27) | -1.20 (4.57) | -1.26 (3.34) | -0.44 (4.67) | 1.09 (3.99) | -1.17 (2.38) | -0.32 (3.48) | 0.72 (3.89) | -2.74 (3.73) | -0.77 (3.78) | 0.37 (3.33) |
| F3 | -1.06 (3.78) | -1.02 (5.13) | 0.53 (4.85) | -0.31 (3.37) | 0.42 (4.50) | 1.02 (3.49) | -0.99 (2.09) | -0.59 (3.20) | -0.05 (2.40) | -1.25 (2.07) | -0.35 (2.49) | 0.22 (2.16) |
| F4 | -1.41 (2.55) | 0.05 (5.29) | 0.71 (4.36) | 0.33 (3.37) | 1.64 (4.35) | 1.59 (3.25) | -0.52 (4.07) | -0.16 (2.90) | -0.35 (3.15) | -0.03 (3.19) | -0.11 (1.69) | -0.11 (2.53) |
| F7 | -2.24 (4.05) | -1.66 (4.35) | 0.87 (5.37) | -0.01 (3.56) | 0.61 (4.71) | 2.84 (4.38) | -0.13 (2.82) | 0.39 (3.32) | 1.82 (3.39) | -0.16 (3.50) | 0.23 (3.23) | 0.98 (2.75) |
| F8 | -0.91 (6.28) | -0.01 (6.77) | 1.85 (6.53) | 0.30 (4.21) | 1.73 (5.97) | 2.58 (4.45) | -0.43 (2.69) | 0.88 (3.91) | 1.17 (3.25) | -1.00 (2.63) | 0.18 (1.96) | 0.38 (1.66) |
| FC5 | -0.55 (4.92) | -1.53 (5.55) | 1.66 (4.77) | -0.50 (4.35) | -0.43 (4.48) | 1.71 (3.73) | -1.33 (2.89) | -1.51 (4.03) | 0.19 (3.28) | -1.55 (3.25) | -0.81 (3.62) | 0.02 (2.84) |
| FC6 | -1.49 (2.22) | -1.04 (6.83) | 1.30 (4.82) | -0.88 (1.90) | -0.68 (3.62) | 1.59 (3.30) | -1.04 (1.62) | -1.19 (2.46) | 0.43 (2.55) | -1.41 (3.19) | -1.13 (3.44) | -0.25 (2.61) |
| T7 | -2.57 (3.79) | -2.11 (4.68) | 0.23 (4.95) | -0.64 (2.81) | 0.60 (4.06) | 2.13 (2.95) | -0.42 (1.98) | 0.35 (2.87) | 0.92 (2.11) | -0.58 (3.27) | 0.82 (4.51) | 0.47 (3.08) |
| T8 | -0.77 (3.46) | 0.25 (5.25) | 2.22 (5.04) | -0.09 (2.96) | 1.58 (4.47) | 3.15 (4.17) | -0.68 (2.28) | 0.58 (3.56) | 1.28 (3.78) | -0.91 (2.51) | 0.95 (3.06) | 0.83 (2.42) |
| P7 | -2.62 (2.84) | -2.30 (4.64) | -0.53 (4.19) | -0.77 (2.33) | 0.23 (3.73) | 1.48 (2.50) | -1.32 (2.25) | -0.89 (3.30) | -0.24 (2.57) | 0.10 (1.54) | 0.69 (2.12) | 0.65 (2.03) |
| P8 | -1.81 (3.66) | -0.72 (7.14) | 1.25 (5.91) | -0.85 (3.17) | 1.09 (5.65) | 2.58 (5.43) | -1.39 (2.47) | 0.29 (4.95) | 0.84 (5.26) | -0.72 (2.15) | 0.97 (5.03) | 1.03 (5.66) |
| O1 | -2.26 (3.80) | -2.48 (4.58) | -0.32 (4.58) | -0.58 (2.17) | -0.05 (2.94) | 1.43 (2.50) | -1.72 (2.85) | -1.66 (3.33) | -0.74 (2.65) | 0.24 (1.79) | 0.48 (2.01) | 0.61 (1.75) |
| O2 | -1.68 (2.86) | -1.43 (5.78) | 0.56 (4.55) | -0.65 (2.22) | 0.39 (3.69) | 1.88 (4.12) | -1.91 (2.59) | -1.13 (3.62) | -0.20 (3.94) | -0.22 (1.88) | 0.70 (3.17) | 1.03 (3.65) |
