## Supplementary material for "BrainPT, a Cognitive Mini-Game App for Short-Form Video Use in Young Adults: Neural and Behavioral Pilot Study": Multimedia_Appendix_4_TREND_checklist

### Multimedia Appendix 4. Completed TREND checklist

Reporting follows the TREND statement for nonrandomized evaluations of behavioral and public health interventions. The checklist applies to Study 2, the nonrandomized controlled comparison; Study 1 is a within-subject laboratory study to which several items do not apply. Section references are to the main manuscript unless stated otherwise.

| Item | TREND item | How it is addressed | Where reported |
|---|---|---|---|
| **Title and Abstract** | | | |
| 1 | Information on how unit were allocated to interventions | Title states the design as a pilot study of a mobile intervention; Methods, "Study Design" states that Study 2 used a nonrandomized nonequivalent control group with assignment by recruitment order and timing. | Title; Methods > Study Design; Methods > Group Allocation (Study 2) |
| 1 | Structured abstract | Abstract is structured as Background, Objective, Methods, Results, Conclusions. | Abstract |
| 1 | Information on target population or study sample | Abstract Methods and Participants give university students in Seoul, with sample sizes for both studies. | Abstract; Methods > Participants |
| **Introduction** | | | |
| 2 | Scientific background and explanation of rationale | Short-form viewing is framed as a neural state; the limits of time-based and self-report outcomes are set out. | Introduction > Background |
| 2 | Theories used in designing behavioral interventions | Top-down/bottom-up attention and prefrontal control theory are cited as the basis for the intervention target. | Introduction > Background [9,10] |
| 3 | Specific objectives and hypotheses | Four hypotheses (H1-H4) are stated. | Introduction > Objectives and Hypotheses |
| **Methods** | | | |
| 4 | Eligibility criteria; method of recruitment; recruitment setting; inclusion/exclusion | Inclusion and exclusion criteria, recruitment route, and setting are given. | Methods > Participants |
| 5 | Details of the interventions intended for each study condition and how/when administered | Threshold, task set, delivery, and adherence logging are described; screens are shown in Figure 1 and Multimedia Appendix 1. | Methods > Intervention (BrainPT); Figure 1; Multimedia Appendix 1 |
| 5 | Content: what was delivered | Ten cognitive tasks, five per triggered set, formats borrowed from the Comprehensive Attention Test. | Methods > Intervention (BrainPT); Multimedia Appendix 1 |
| 5 | Delivery method | iOS application distributed via TestFlight; launches when the cumulative daily short-form threshold (30 min) is reached. | Methods > Intervention (BrainPT) |
| 5 | Unit of delivery / unit of assignment | Delivered and assigned at the level of the individual participant. | Methods > Group Allocation (Study 2) |
| 5 | Deliverer, setting, exposure quantity and duration, time span | Self-administered at home over a 14-day window preceding visit 2; exposure quantified from app logs. | Methods > Procedure; Methods > Measures |
| 5 | Activities to increase compliance or adherence | Adherence was verified objectively from trial counts, accuracy, and daily session counts; no incentive was tied to adherence. | Methods > Intervention (BrainPT) |
| 6 | Clearly defined primary and secondary outcome measures | Daily short-form viewing time is the primary outcome; ASRS total and neural indices are secondary. | Methods > Outcomes |
| 6 | Methods used to collect data and any quality-control methods | EEG, fNIRS, app logs, and self-report instruments with their processing and quality criteria. | Methods > Measures; Methods > Neuroimaging Data Processing; Multimedia Appendix 2 |
| 7 | Sample size determination and, when applicable, | No a priori power calculation; sample size was determined by feasibility. Minimum detectable effect is reported instead. | |

| | | | |
|---|---|---|---|
| | explanation of interim analyses and stopping rules | | Methods > Statistical Analysis; Discussion > Limitations |
| 8 | Unit of assignment; method used to assign units; inclusion of any aspect of blinding | Assignment was by recruitment order and timing, not randomized. Participants were not blinded; outcome assessors were not blinded. | Methods > Group Allocation (Study 2) |
| 9 | Description of smallest unit being analyzed | The individual participant. | Methods > Statistical Analysis |
| 10 | Statistical methods used to compare study groups | Change-score t tests and analysis of covariance with the visit-1 value as covariate; Welch t tests for unequal variances. | Methods > Statistical Analysis |
| 10 | Statistical methods used for additional analyses | Benjamini-Hochberg false-discovery-rate control, equivalence testing, cluster-based correction, per-protocol and sensitivity analyses. | Methods > Statistical Analysis |
| 10 | Methods for imputing missing data | No imputation. Indices were analyzed on the largest analyzable sample and the sample is stated for each. | Methods > Statistical Analysis; Multimedia Appendix 2 |
| **Results** | | | |
| 11 | Flow of participants through each stage | Enrollment, allocation, exclusions with reasons, and analyzed numbers are given for both studies. | Results > Study 1; Results > Study 2; Table 1 |
| 12 | Dates defining the periods of recruitment and follow-up | Study periods and inter-visit intervals are reported. | Methods > Procedure; Table 1 |
| 13 | Baseline demographic and clinical characteristics of participants in each condition | Reported by group with baseline comparisons. | Table 1 |
| 13 | Baseline comparisons of those lost to follow-up and those retained | Excluded participants and the reasons are reported; the per-protocol analysis excluding one participant is also reported. | Results > Study 2; Discussion > Limitations |
| 14 | Number of participants assigned to each group and number analyzed | As-assigned and analyzed numbers are given separately for behavioral and neural indices. | Results > Study 2; Table 1 |
| 15 | Data on estimated effect and precision | Effect sizes with 95% CIs are reported for all primary comparisons. | Results (throughout) |
| 16 | Summary results of any other analyses performed | Equivalence testing, exploratory connectivity, and regional block contrasts are reported. | Results > Neural Carry-Over; Results > Exploratory Connectivity Analysis; Multimedia Appendix 2 |
| 17 | Summary of important adverse events or unintended effects | No adverse events were observed or reported. | Results > Study 2 |
| **Discussion** | | | |
| 18 | Interpretation of the results | Results are interpreted against the stated hypotheses. | Discussion > Principal Results |
| 19 | Generalizability of the trial findings | Generalization is limited to the sampled population and the 14-day exposure. | Discussion > Limitations |
| 20 | Overall interpretation of the results in the context of current evidence | Findings are placed against prior intervention and cognitive-training literature. | Discussion > Comparison With Prior Work |

Reference: Des Jarlais DC, Lyles C, Crepaz N; TREND Group. Improving the reporting quality of nonrandomized evaluations of behavioral and public health interventions: the TREND statement. Am J Public Health. 2004;94(3):361-366. [doi: 10.2105/ajph.94.3.361] [Medline: 14998794]
