## Supplementary material for "BrainPT, a Cognitive Mini-Game App for Short-Form Video Use in Young Adults: Neural and Behavioral Pilot Study": Multimedia_Appendix_5_CONSORT_EHEALTH

### Multimedia Appendix 5. Completed CONSORT-EHEALTH checklist (V1.6.1)

CONSORT-EHEALTH is required for randomized trials of eHealth and mHealth interventions. Study 2 of this report is a nonrandomized controlled pretest–posttest comparison, so the primary reporting guideline is the TREND statement (Multimedia Appendix 4). The CONSORT-EHEALTH form is completed here in addition, because its intervention-description items (item 5 and its subitems in particular) are the fullest available account of what an eHealth intervention was and how it was used, and because JMIR readers expect them. Items that depend on randomization (8a, 8b, 9) are marked not applicable; item 10 is answered by describing how assignment was actually made. Section references are to the main manuscript unless stated otherwise.

| Item | CONSORT-EHEALTH item | How it is addressed | Where reported |
|---|---|---|---|
| **Title and Abstract** | | | |
| 1a-i | Identify the mode of delivery in the title | The title names the delivery mode: "BrainPT, a Cognitive Mini-Game App for Short-Form Video Use in Young Adults: Neural and Behavioral Pilot Study." | Title |
| 1a-ii | Non-web-based components or important co-interventions in title | There were no co-interventions. The laboratory neuroimaging sessions are measurement, not intervention, and are signalled by "Neural and Behavioral" in the title. | Title |
| 1a-iii | Primary condition or target group in the title | The target group ("Young Adults") and the target behavior ("Short-Form Video Use") both appear in the title. | Title |
| 1b-i | Key features/functionalities/ components of the intervention and comparator in the METHODS section of the ABSTRACT | The abstract Methods state that the experimental group used BrainPT daily and that the control group did not, under the same in-laboratory protocol at both visits. | Abstract > Methods |
| 1b-ii | Level of human involvement in the METHODS section of the ABSTRACT | BrainPT is fully automated: the app triggers on a device-side time threshold and requires no human facilitator. The abstract describes daily unsupervised at-home use. | Abstract > Methods |
| 1b-iii | Open vs. closed, web-based vs. face-to-face assessments in the METHODS section of the ABSTRACT | The abstract states that neural measurement took place in the laboratory at both visits and that usage logs and self-report indices were collected over the intervention period. | Abstract > Methods |
| 1b-iv | RESULTS section in abstract must contain use data | The abstract reports the primary behavioral outcome as means for both groups (143.5 to 50.9 vs 104.8 to 132.9 min/day). Adherence figures (median 372 trials over 13 days) are in Table 1 rather than the abstract, for the 450-word limit. | Abstract > Results; Table 1 |
| 1b-v | CONCLUSIONS/DISCUSSION section of abstract for negative trials | The neural carry-over outcome was null and is stated as such: "no neural index separated the groups after 2 weeks," with the conclusion noting the small, nonrandomized samples and the need for a randomized trial with objectively logged use. | Abstract > Results; Abstract > Conclusions |
| **Introduction** | | | |
| 2a-i | Problem and the type of system/ solution | Short-form viewing is framed as a stimulus-driven neural state, and existing interventions are described as targeting time of use alone without showing what changed. The proposed solution is an insertion-type mobile intervention. | Introduction > Background |
| 2a-ii | Scientific background, rationale: What is known about the (type of) system | Prior behavioral, fMRI, EEG, and fNIRS findings on short-form use are reviewed, together with evidence that cognitive-training and smartphone-reduction interventions change trained tasks but transfer poorly. | Introduction > Background [2-9] |
| 2b | Specific objectives or hypotheses | Two objectives and four hypotheses (H1-H4) are stated explicitly, with H1-H3 neuroimaging and H4 behavioral. | Introduction > Objectives and Hypotheses; Textbox 1 |
| **Methods: Trial Design** | | | |

| | | | |
|---|---|---|---|
| 3a | Description of trial design including allocation ratio | Study 1 is a within-subject repeated-measures design (two visits, no app use between). Study 2 is a nonrandomized controlled pretest-posttest design with a nonequivalent control group; 15 were assigned to the experimental and 8 to the control condition by recruitment route and timing. | Methods > Study Design; Methods > Group Allocation (Study 2) |
| 3b-i | Bug fixes, downtimes, content changes | No content change, version update, or feature change was made to the app during the intervention period. No downtime was reported by participants or observed in the upload logs. Task set and difficulty were fixed for all participants throughout. | Methods > Intervention (BrainPT) |
| **Methods: Participants** | | | |
| 4a-i | Computer / Internet literacy | All participants were university students in their 20s-30s who were daily short-form viewers on their own smartphones; no separate literacy screen was applied because habitual smartphone use was itself an eligibility criterion. | Methods > Participants |
| 4a-ii | Open vs. closed, web-based vs. face-to-face assessments | Recruitment was open, through public notices on a university-community application. Eligibility was confirmed by a closed pre-survey. Neural outcomes were assessed face-to-face in the laboratory; usage and self-report outcomes were collected through a closed online form open only to enrolled participants. | Methods > Participants; Methods > Measures |
| 4a-iii | Information given during recruitment | The recruitment notice stated the purpose, the two laboratory visits, the approximate one-hour duration per visit, the honorarium, and the eligibility and exclusion criteria. Experimental and control conditions were advertised through separate notices, so applicants knew the condition of the notice they answered but could not choose between conditions. | Methods > Participants; Methods > Group Allocation (Study 2) |
| 4b-i | Report if outcomes were (self-)assessed through online questionnaires | Daily short-form viewing time was self-reported through an online form and cross-checked against a submitted device screen-time screenshot; ASRS and the five daily self-report items were also collected online. This is stated as a limitation. | Methods > Measures > Surveys; Discussion > Limitations > Measurement |
| 4b-ii | Report how institutional affiliations are displayed | The recruitment notice and consent form identified the Department of Psychology, Yonsei University as the conducting institution. The app itself carried no institutional branding beyond the study name. | Methods > Ethical Considerations |
| **Methods: Interventions** | | | |
| 5-i | Mention names, credentials, affiliations of the developers, sponsors, and owners | BrainPT was developed by the authors, with SMH leading the implementation; all authors are affiliated with the Department of Psychology, Yonsei University. The funders had no role in design, analysis, or reporting. | Conflicts of Interest; Authors' Contributions; Funding Statement |
| 5-ii | Describe the history/development process | The intervention concept was developed in a student cognitive science society forum and implemented as an iOS application by the author team; task formats were adapted from the Comprehensive Attention Test to target the functions reported to be weakened by short-form use. | Methods > Intervention (BrainPT); Acknowledgments |
| 5-iii | Revisions and updating | The version used was fixed for the whole intervention period; no revision or update was pushed to participants during the study. | Methods > Intervention (BrainPT) |
| 5-iv | Quality assurance methods | Trial count, accuracy, and response time were logged per game and uploaded automatically, which allowed unsupervised use to be verified objectively. Logs were inspected for every participant; one participant whose logs | Methods > Intervention (BrainPT); Methods > Statistical Analysis |

| | | | |
|---|---|---|---|
| | | showed only an in-laboratory exposure is reported separately in a per-protocol analysis. | |
| 5-v | Ensure replicability by publishing the source code, and/or providing screenshots/screen-capture video, and/or providing flowcharts of the algorithms used | Figure 1 shows the app; Multimedia Appendix 1 gives a screen and a written rule for each of the 10 tasks, together with the intended cognitive function and the set structure. The trigger rule (cumulative daily short-form time of 30 minutes) and the fixed-difficulty policy are stated. | Figure 1; Multimedia Appendix 1; Methods > Intervention (BrainPT) |
| 5-vi | Digital preservation | The application was distributed through iOS TestFlight for research use only and is not commercially available. Build and task specifications are retained by the corresponding author; Multimedia Appendix 1 preserves the task content in the published record. | Conflicts of Interest; Data Availability; Multimedia Appendix 1 |
| 5-vii | Access | Access was restricted to enrolled experimental-group participants through TestFlight invitation. The app is free, carries no advertising, and no revenue is received from it. | Methods > Intervention (BrainPT); Conflicts of Interest |
| 5-viii | Mode of delivery, features/ functionalities/components of the intervention and comparator, and the theoretical framework | Delivery is a self-contained iOS app on the participant's own phone. When cumulative daily short-form time reaches 30 minutes the app launches, presents one set of 5 of 10 cognitive tasks, and closes when the set is complete; viewing is interrupted but not blocked afterward. The theoretical framework is the top-down/bottom-up account of attention and prefrontal cognitive control. The comparator received no app and completed the same daily self-reports. | Methods > Intervention (BrainPT); Introduction > Background [10,11] |
| 5-ix | Describe use parameters | Use was quantified as total trials, accuracy, daily session count, and first-use date over the 14-day window preceding visit 2. Experimental participants completed 12 to 628 trials (median 372) over a median of 13 days; control participants had 12 to 13 trials over 1 to 2 days, all near their own visit-2 session. | Methods > Group Allocation (Study 2); Table 1 |
| 5-x | Clarify the level of human involvement | None during the intervention period. The app is fully automated and no coaching, monitoring feedback, or contact from the research team took place between visits. | Methods > Intervention (BrainPT); Methods > Procedure > Study 2 |
| 5-xi | Report any prompts/reminders used | The only prompt was the app's own automatic launch at the 30-minute daily threshold. No push reminders, emails, or messages were sent to encourage app use or self-report completion. | Methods > Intervention (BrainPT) |
| 5-xii | Describe any co-interventions (incl. training/support) | None. Both groups completed the same daily self-report and the same two laboratory visits; the experimental group additionally installed the app after visit 1. | Methods > Procedure > Study 2 |
| **Methods: Outcomes** | | | |
| 6a-i | Online questionnaires: describe if they were validated for online use and apply CHERRIES items to describe how the questionnaire was designed/deployed | The ASRS is a published 18-item instrument, administered here in its standard item set with each item scored 0 to 4. The five daily items were purpose-written for this study and are quoted in full in the Methods. Forms were open only to enrolled participants, one submission per participant per day, with no incentive tied to completion. | Methods > Measures > Surveys; Methods > Measures > Daily Self-Report [23] |
| 6a-ii | Describe whether and how "use" (including intensity of use/ dosage) was defined/measured/ monitored | Use was defined as app-logged activity in the 14-day window preceding visit 2 and monitored from automatically uploaded per-game records (trial count, accuracy, response time, session date). Dose is reported as total trials and days of use, and a dose-response analysis by baseline use is reported as exploratory. | Methods > Intervention (BrainPT); Methods > Outcomes; Table 1 |
| 6a-iii | Describe how qualitative feedback from participants was obtained | Structured daily self-report items on sustained attention, emotional stability, impulse inhibition, plan execution, and overall satisfaction were collected throughout the | Methods > Measures > Surveys; Results > Behavioral Change After 2 Weeks of Use |

| | | | |
|---|---|---|---|
| | | intervention period. At visit 2 the experimental group additionally completed three fixed-choice post-survey items on the perceived effect of the app and two open-ended items inviting free comment on the experience of use and on anything they found inconvenient or would want improved; these are reported descriptively in the Results. | |
| 6b | Any changes to trial outcomes after the trial commenced | The outcome hierarchy was fixed before analysis and is stated in Methods. Two Study 2 analyses use visit-1 data only and are presented as pre-intervention state description; the connectivity analysis outside the pre-specified channel pairs and the dose-response analysis were not pre-specified and are labelled exploratory throughout. | Methods > Outcomes; Results > Exploratory Connectivity Analysis |
| **Methods: Sample Size, Randomization, Blinding** | | | |
| 7a-i | Describe whether and how expected attrition was taken into account when calculating the sample size | No a priori power calculation was performed; the sample size was set by feasibility for a pilot study. The minimum detectable effect is reported instead, with the observation that detecting d=0.5 would require about 64 participants per group. | Methods > Statistical Analysis; Discussion > Limitations > Sample and Power |
| 7b | Explanation of any interim analyses and stopping guidelines | No interim analysis was conducted and no stopping rule was applied. | Methods > Statistical Analysis |
| 8a | Method used to generate the random allocation sequence | Not applicable — this is a nonrandomized controlled study. | Methods > Group Allocation (Study 2) |
| 8b | Type of randomisation; details of any restriction | Not applicable — this is a nonrandomized controlled study. | Methods > Group Allocation (Study 2) |
| 9 | Mechanism used to implement the random allocation sequence | Not applicable — this is a nonrandomized controlled study. | Methods > Group Allocation (Study 2) |
| 10 | Who generated the allocation sequence, who enrolled participants, and who assigned participants | Assignment was by recruitment route and timing rather than by a sequence: the experimental notice was posted and that group measured first, after which the control notice was posted. Enrollment and assignment were carried out by the study team; applicants could not choose between conditions. | Methods > Group Allocation (Study 2) |
| 11a-i | Specify who was blinded, and who wasn't | Nobody was blinded. Because each applicant answered a notice that described its own condition, participants knew their condition, and the experimenters who ran the sessions and processed the data were the same team. | Methods > Group Allocation (Study 2) |
| 11a-ii | Discuss whether participants knew which intervention was the "intervention of interest" and which one was the "comparator" | Participants in the experimental group knew they were receiving the intervention of interest; control participants knew they were completing self-reports only. Expectancy effects on the self-reported primary outcome therefore cannot be excluded and are discussed as a limitation. | Methods > Group Allocation (Study 2); Discussion > Limitations |
| 11b | Description of the similarity of interventions | The two conditions were not matched: the control group received no app and no active comparator. Both groups completed identical laboratory protocols at both visits and the identical daily self-report. | Methods > Procedure > Study 2 |
| 12a-i | Imputation techniques to deal with attrition / missing values | No imputation was performed. Each index was analyzed on the largest analyzable sample, the sample size is stated for every comparison, and a sensitivity analysis restricting behavioral outcomes to the neural-analysis sample is reported. | Methods > Statistical Analysis; Multimedia Appendix 2 |
| 12b | Methods for additional analyses, such as subgroup analyses and adjusted analyses | Analysis of covariance with the visit-1 value as covariate was run alongside change-score t tests, with pre-specified treatment of disagreement between them. Multiplicity was controlled with the Benjamini-Hochberg false-discovery rate; absences of effect were addressed with equivalence | Methods > Statistical Analysis |

| | | | |
|---|---|---|---|
| | | testing; the higher-resolution spectral analysis used cluster-based correction; band-power comparisons additionally controlled for the aperiodic component and residual artifact. | |
| **Results** | | | |
| 13a | Numbers of participants assigned, receiving intended treatment, and analyzed for the primary outcome | Study 1 enrolled 15 and analyzed 9. Study 2 assigned 23 (15 experimental, 8 control) and analyzed 21 for the behavioral primary outcome (13 experimental, 8 control) and 18 for neural indices. | Results; Table 1 |
| 13b-i | Attrition diagram | Stage-by-stage counts and exclusion reasons are tabulated rather than drawn, in the Sample block of Table 1 and its footnote e. | Table 1 |
| 14a-i | Indicate if critical "secular events" fell into the study period | No platform policy change, public campaign, or comparable event affecting short-form use is known to have occurred during either recording period (Study 1, January to February 2025; Study 2, September to November 2025). Measurement timing nonetheless differed systematically between groups and is reported and discussed. | Methods > Participants; Table 1; Discussion > Limitations > Nonrandomized Assignment |
| 14b | Why the trial ended or was stopped | Recruitment ended when the planned feasibility sample was reached; the study was not stopped early. | Methods > Participants |
| 15-i | Report demographics associated with digital divide issues | Age, sex, baseline daily short-form use, and ASRS total are reported by group with between-group comparisons. All participants were university students who owned and habitually used a smartphone, so the sample does not span a digital divide; this narrows generalizability and is stated. | Table 1; Discussion > Limitations |
| 16-i | Report multiple "denominators" and provide definitions | Assigned, behavioral-analysis, EEG-analysis, and fNIRS-analysis samples are reported separately for each group in Table 1, and the relevant n is given with every comparison in the Results. | Table 1; Results |
| 16-ii | Primary analysis should be intent-to-treat | The primary behavioral analysis is as-assigned, including the participant whose logs indicated only an in-laboratory exposure. A per-protocol analysis excluding that participant is reported alongside it. | Methods > Statistical Analysis; Results > Behavioral Change After 2 Weeks of Use |
| 17a-i | Presentation of process outcomes such as measures of use and intensity of use | Total app trials (median and range) and days of use are reported by group in Table 1, with the control group's single in-laboratory exposure described explicitly. | Table 1; Methods > Group Allocation (Study 2) |
| 18-i | Subgroup analysis of users only | The per-protocol analysis, which excludes the participant without genuine at-home use, serves this role. A dose-response analysis by baseline use is reported as exploratory. | Results > Behavioral Change After 2 Weeks of Use |
| 19-i | Include privacy breaches, technical problems | No privacy breach, data loss, or app crash was reported or observed. Recording-quality failures and missing block markers affected neural data collection and are itemized as exclusions. | Results; Table 1, footnote e |
| 19-ii | Include qualitative feedback from participants or observations from staff/researchers | No adverse event or unintended effect was reported by participants or observed by the research team. Free-text responses did report unintended burden: several participants described the inserted task as a nuisance they came to avoid, one reported that the screen-time trigger fired during non-short-form use of the same app, and the most common improvement request was for greater task variety. | Results > Behavioral Change After 2 Weeks of Use; Discussion |
| **Discussion** | | | |
| 20-i | Typical limitations in ehealth trials | The stated limitations are nonrandomized assignment confounded with measurement timing, small per-group samples with the corresponding minimum detectable effect, reliance on self-report and submitted screenshots rather than | Discussion > Limitations |

| | | | |
|---|---|---|---|
| | | direct app-level metering for the primary outcome, and sensor-space EEG constraints on the connectivity claims. | |
| 21-i | Generalizability to other populations | Findings are limited to young adult university students in Seoul who were already motivated to reduce their own use; extension to other ages, to non-help-seeking users, and to Android is not established. | Discussion > Limitations |
| 21-ii | Discuss if there were elements in the RCT that would be different in a routine application setting | Participation involved two laboratory visits, a daily self-report, and an honorarium, none of which exist in routine use; observed adherence should therefore not be read as adherence outside a study. Conversely, the 30-minute threshold was fixed for everyone, whereas a deployed version would plausibly be user-set. | Discussion > Limitations; Discussion > Principal Results |
| 22-i | Restate study questions and summarize the answers | The Conclusions restate both questions and answer them: a single insertion moved the prefrontal state toward the goal-directed pole, and 2 weeks of use was accompanied by a 65% fall in daily viewing, while no neural index separated the groups after 2 weeks. | Discussion > Principal Results; Conclusions |
| 22-ii | Highlight unanswered new questions, suggest future research | A randomized trial with objectively logged use is identified as the required next step, together with a longer exposure to test whether resting-state change emerges and pre-specified testing of the posterior beta-band coupling seen here only as exploratory. | Discussion > Limitations > Duration; Discussion > Limitations > Exploratory Findings; Conclusions |
| **Other Information** | | | |
| 23 | Registration number and name of trial registry | Not registered. Assignment was not randomized, so the study does not meet the definition of a clinical trial requiring prospective registration; this is stated in the Methods. | Methods > Study Design |
| 24 | Where the full trial protocol can be accessed | No separate protocol document is published. The full procedure, measures, and analysis plan are given in the Methods and Multimedia Appendices 1 to 3. | Methods; Multimedia Appendices 1-3 |
| 25 | Sources of funding and other support; role of funders | Supported by the Institute for Higher Education Innovation, Yonsei University (KRW 6,500,000) and the Institute for Global Engagement and Empowerment, Yonsei University (KRW 3,500,000). The funders had no role in study design, data collection and analysis, the decision to publish, or preparation of the manuscript. | Funding Statement |
| X26-i | Comment on ethics committee approval | Approved by the Institutional Review Board of the Department of Psychology, Yonsei University (7001988-202509-HR-2369-04), and conducted in accordance with the Declaration of Helsinki. | Methods > Ethical Considerations |
| X26-ii | Outline informed consent procedures | Before starting, all participants were informed of the purpose, procedures, anticipated discomforts, scope of data use, and their right to withdraw at any time without penalty, and gave written consent. | Methods > Ethical Considerations |
| X26-iii | Safety and security procedures | App-usage records were transmitted to a private repository accessible only to the research team. Data were recorded with participant initials and a study number only, with no name or contact information stored; initials will be replaced with random identifiers for any public release. Raw EEG and fNIRS recordings are not publicly deposited because signal characteristics are potentially identifying. | Methods > Ethical Considerations; Data Availability |
| X27-i | State the relation of the study team towards the system being evaluated | The BrainPT application was developed by the authors and SMH led its implementation, so the authors hold a nonfinancial interest in the intervention under evaluation. The app is not commercially available, was distributed only | Conflicts of Interest |

through TestFlight for research use, and the authors receive no revenue and hold no patent or equity related to it.

---

Reference: Eysenbach G; CONSORT-EHEALTH Group. CONSORT-EHEALTH: improving and standardizing evaluation reports of Web-based and mobile health interventions. J Med Internet Res. 2011;13(4):e126. [doi: 10.2196/jmir.1923] [Medline: 22209829]
